# A novel intronic GAA repeat expansion in *FGF14* causes autosomal dominant adult-onset ataxia (SCA50, ATX-FGF14)

**DOI:** 10.1101/2022.10.21.22281020

**Authors:** Haloom Rafehi, Justin Read, David J Szmulewicz, Kayli C. Davies, Penny Snell, Liam G Fearnley, Liam Scott, Mirja Thomsen, Greta Gillies, Kate Pope, Mark F Bennett, Jacob E Munro, Kathie J. Ngo, Luke Chen, Mathew J Wallis, Ernest G Butler, Kishore R Kumar, Kathy HC Wu, Susan E Tomlinson, Stephen Tisch, Abhishek Malhotra, Matthew Lee-Archer, Egor Dolzhenko, Michael A. Eberle, Leslie J Roberts, Brent L Fogel, Norbert Brüggemann, Katja Lohmann, Martin B. Delatycki, Melanie Bahlo, Paul J Lockhart

**Author notes:** These authors contributed equally to this work (Haloom Rafehi, Justin Read, David J Szmulewicz). These authors contributed equally to this work (Melanie Bahlo, Paul J Lockhart). Correspondence to Prof Melanie Bahlo, The Walter and Eliza Hall Institute of Medical Research, 1G Royal Parade, Parkville, Victoria 3052, Australia; and Prof Paul Lockhart, Murdoch Children’s Research Institute, Flemington Rd, Parkville, Victoria 3052, Australia.

## Abstract

Adult-onset cerebellar ataxias are a group of neurodegenerative conditions that challenge both genetic discovery and molecular diagnosis. In this study, we identified a novel intronic GAA repeat expansion in the gene encoding Fibroblast Growth Factor 14 (*FGF14*). Genetic analysis identified 4/95 previously unresolved Australian affected individuals (4.2%) with (GAA)_>335_ and a further nine individuals with (GAA)_>250_. Notably, PCR and long-read sequence analysis revealed these were pure GAA repeats. In comparison, no controls had (GAA)_>300_ and only 2/311 control individuals (0.6%) encoded a pure (GAA)_>250_. In a German validation cohort 9/104 (8.7%) of affected individuals had (GAA)_>335_ and a further six had (GAA)_>250_. In comparison no controls had (GAA)_>335_ and 10/190 (5.3%) encoded (GAA)_>250_. The combined data suggests (GAA)_>335_ are disease-causing and fully penetrant [P-value 6.0×10^−8^, OR 72 (95% CI=4.3-1227)], while (GAA)_>250_ is likely pathogenic, albeit with reduced penetrance. Affected individuals had an adult-onset, slowly progressive cerebellar ataxia with a clinical phenotype that may include vestibular impairment, hyper-reflexia and autonomic dysfunction. A negative correlation between age at onset and repeat length was observed (R2=0.44 p=0.00045, slope = -0.12). This study demonstrates the power of genome sequencing and advanced bioinformatic tools to identify novel repeat expansion loci via model free, genome-wide analysis and identifies SCA50/ATX-FGF14 is a frequent cause of adult-onset ataxia.

## INTRODUCTION

Spinocerebellar ataxias (SCAs) are a heterogenous group of progressive neurological disorders that are estimated to affect 1:33,000 individuals.^1^ SCAs are caused by pathogenic expansions of short tandem repeats (STRs, also called microsatellites), in addition to deleterious point-mutations. STRs are repeated nucleotide motifs of 2-6 base pairs (bp) that are known contributors to genetic polymorphism. There are ∼239,000 STRs documented in the UCSC Genome Browser^2^ and although current catalogues are incomplete^3^ STR loci are unstable and can rapidly mutate to alternative motifs or alter in length. About 40 STRs have been shown to be pathogenic if they expand beyond locus-specific thresholds and result in repeat expansion (RE) disorders.^4^ These include more common causes of ataxia, such as autosomal dominant SCA 1, 2, 3, 6 and 7, and autosomal recessive Friedreich ataxia, as well as rarer forms such as SCA36 and 37. Many people presenting with ataxia receive a clinical diagnosis of idiopathic late onset cerebellar ataxia (ILOCA)^5^, or sporadic adult-onset ataxia (SAOA).^6^ Ataxia may present with other clinical features, including neuropathy, vestibular dysfunction, parkinsonism, cognitive impairment and psychiatric features, which may provide clues to an underlying genetic diagnosis. However, despite routine clinical testing for the more common causes of ataxia, only ∼10-30% of individuals with a clinical diagnosis of ataxia receive a genetic diagnosis.^7^ This is thought to be in part due to as yet unidentified genetic causes, including REs, likely located in non-coding regions of the genome.^4^

The identification of pathogenic REs has historically been difficult. However, discovery and detection of complex genetic variants, such as REs, in genome sequencing data have improved due to the advances in bioinformatic tools available to identify both known and novel RE.^8,9^ Using these tools, we recently identified the RE that causes cerebellar ataxia with neuropathy and bilateral vestibular areflexia syndrome (CANVAS), a common cause of adult-onset cerebellar ataxia.^10^ This RE, a novel (AAGGG)_n_ RE in *RFC1*, had previously evaded discovery despite presenting a well-defined recessive disease phenotype and a strong linkage region.

In this study we tested the hypothesis that advanced bioinformatic tools and second and third generation sequencing technologies provide an opportunity to perform model-free discovery of novel pathogenic REs in cohorts of unrelated individuals with overlapping but heterogeneous disorders. We identified a novel (GAA)_n_ RE within intron 1 of *FGF14* that appears to represent the most common genetic cause of adult-onset ataxia described to date.

## MATERIALS AND METHODS

### Cohort recruitment and clinical phenotyping

The Royal Children’s Hospital Human Research Ethics Committee (HREC 28097) and the Walter and Eliza Hall Institute of Medical Research Human Research Ethics Committee (HREC 18/06) approved the study. The study ID codes utilized in this research meet the requirements of the Ethics Committees regarding de-identification and are known only to the research group. Individuals with suspected genetic cerebellar ataxia were recruited from multiple clinicians in Australia. Informed consent was obtained from all 95 participants and details were collected from clinical assessments and review of medical records. All participants were examined clinically by a consultant neurologist, with eight assessed by sub-specialist neuro-otologists. Cerebellar functional assessment included evaluation for gait ataxia, cerebellar dysarthria and four limb appendicular ataxia. Oculomotor assessment included documentation (in most) of any abnormal nystagmus, visual pursuit and saccades to target. Brain MRI scans were assessed visually for regional cerebellar atrophy. Eight individuals underwent nerve conduction studies, one of these had only lower limb studies. The other seven had motor studies of the median, ulnar, peroneal and tibial nerves, including F-waves. Six of these seven participants also had upper limb sensory studies including antidromic and orthodromic median, ulnar and radial digital studies while one person had limited upper limb sensory studies. All eight participants had antidromic sural and superficial peroneal sensory studies. Small nerve fibre studies were performed in six of the eight, with five having QSART testing and six having cutaneous silent period (CSP) studies. The QSART tests a population of C-fibres and the CSP studies test somatic A**δ** fibres. Tilt table testing was undertaken in order to assess autonomic nervous system (ANS) function in four of the eight individuals.

The control cohort was comprised of 215 unrelated adult individuals recruited from multiple sites in Australia, all healthy at the time of blood sampling (22-66 years of age). In addition, a panel of 96 gDNA samples derived from UK Caucasian control individuals, all healthy at the time of DNA sampling (24-65 years of age), were analyzed (HRC-1 control panel, Sigma Aldrich, #06041301).

A validation cohort of 104 unrelated individuals with ataxia of an unidentified cause and 190 control individuals from Lübeck, Germany, were included. The Ethics committee of the University of Lübeck approved clinical assessments and genetic testing of this cohort (AZ16-039). The participants were recruited from the ataxia and vertigo outpatient clinic at the Department of Neurology, University Medical Center Schleswig-Holstein, Campus Lübeck. Vestibulo-oculography, calorimetry, NCS and MRI were performed in a clinical setup. Some of the clinical MRI scans were not available for a personal review. In this case, information was derived from written reports.

### Next generation sequence analysis

Genomic DNA was isolated from peripheral blood and genome sequencing was performed with the TruSeq PCR-free DNA HT Library Preparation Kit and sequenced on the Illumina NovaSeq 6000 platform. GS data generated in-house from 210 unrelated control individuals with no clinical evidence of ataxia was used for genetic studies. Targeted long-read sequencing of *FGF14* was performed using adaptive sampling on an Oxford Nanopore Technologies MinION Mk1B sequencer. Blood derived genomic DNA was used to construct sequencing libraries using the manufacturer’s specifications including the NEBNext Companion Module (New England BioLabs) and SQK-LSK110 Ligation Sequencing Kit (Oxford Nanopore Technologies). FLO-MIN106D flow cells (Oxford Nanopore Technologies) were loaded with library and run for 24 hours before being washed using the Flow Cell Wash Kit (EXP-WSH004, Oxford Nanopore Technologies) and reloaded with additional library for a further 24 hours. Sequencing with adaptive sampling was performed by Readfish (v.0.0.2)^11^, with the entire gene and a 100kb flanking region selected for adaptive sequencing enrichment (hg38 chr13:101610804-102502457).

### Alignment and variant calling and STR analysis

For short-read data, alignment was performed based on the GATK best practice latepipeline.^12,13^ Fastq files were aligned to the hg38 reference genome using BWA-mem, then duplicate marking, local realignment, and recalibration was performed with GATK. The RE detection tools exSTRa^14^ (version 1.1.0) and ExpansionHunter^15^ (version 5.0.0) were used to screen for known pathogenic REs using default parameters. A database of pre-defined pathogenic REs was used (exSTRa in Web resources, file version committed on Jun 26, 2020). These tools utilize paired-end reads to detect expanded STRs: exSTRa uses an empirical cumulative distribution function (ECDF) to determine outliers, while ExpansionHunter estimates the number of repeats in the STR on each allele. Novel RE discovery was performed using ExpansionHunter Denovo^9^ (EHDN, version 0.9.0) using the outlier method (max-irr-mapq=60), comparing 47 individuals with ataxia to 210 controls. EHDN was also used, using default parameters, to profile genome sequence data from the 1000 Genomes Project.^16^ Analysis was performed on the publicly available ‘1000 Genomes 30x on GRCh38’ data collection.^17^ Sample metadata, including pedigree information and ethnicity, was obtained from the metadata provided by the 1000 Genomes Project.^16^ Related individuals were removed from the analysis, leaving 2483 individuals from diverse ethnic backgrounds. We used nf-cavalier, an in-house Nextflow Pipeline^18^, for screening of SNP and indel variants (cavalier in Web resources). The pipeline utilizes GATK variant calls^19^, bcftools^20^, IGV^21^ and ensembl VEP^22^ annotations. Variants were filtered according to the following criteria: minimum VEP impact “MODERATE”; maximum cohort allele frequency 0.3; and present on a curated ataxia gene list. Candidate genes were selected from published ataxia gene panels curated by PanelApp Australia and PanelApp UK. Additional genes were obtained from OMIM with a phenotype referencing ataxia. Dominant, recessive, and compound heterozygous inheritance patterns were further filtered using VEP provided gnomAD allele frequencies (AF) as follows: AF dominant <= 0.0001, AF recessive <= 0.01, AF compound heterozygous <= 0.01. IGV screenshots of putative variant calls were then reviewed manually. For the long-read data, base-calling was performed using the Guppy basecaller with a super-accuracy base-calling model (version 6.2.3, Oxford Nanopore Technologies). STR analysis was performed using tandem-genotypes^23^ (version 1.90) after alignment with LAST (v1409). A common ancestral haplotype was determined for three Australian participants with (GAA)_>250_ by comparing variant sharing around the *FGF14* STR for variations with an gnomAD AF<0.1.

### Molecular genetic studies

We designed a long-range PCR (LR-PCR) assay to test the size of the *FGF14* STR utilizing Phusion Flash High-Fidelity PCR Master Mix (ThermoFisher Scientific, F548). The primers (Table S1) flank the STR and are predicted to amplify a 315bp fragment based on hg38 reference sequence, which includes 50 GAA repeats. PCR was performed in a 20µl reaction with 50 ng genomic DNA and 0.1 µM of forward and reverse primers. A standard 35 cycle PCR protocol was utilized (98°C denaturation for 1 s, 63°C anneal for 10 s, and 72°C extension for 2 min). Products were visualized using agarose gel electrophoresis and fragment analysis of FAM labelled PCR products was performed on capillary array (ABI3730xl DNA Analyzer, Applied Biosystems) and visualized using PeakScanner 2 (Applied Biosystems). The presence of an expanded *FGF14* RE was tested by repeat-primed PCR (RP-PCR) utilizing three primers; FGF14_RPP_ F1_FAM (0.1 µM), FGF14_RPP_AAG_RE_R1 (0.2 µM) and RPP_M13R (0.1 µM, Table S1), with the cycling conditions and analysis described above.

### Genetic analysis of the validation cohort

The validation cohort consisted of 104 unrelated individuals with ataxia, 190 controls, and two healthy parents of one affected individual. All samples were collected at the University of Lübeck and were predominantly of German ethnicity. LR-PCR was applied to these 296 individuals with the primers listed in Table S1 using a standard PCR protocol with an extension time of 2 minutes. Products were separated on an automated sequencing machine (Genetic Analyzer 3500XL, Applied Biosystems) using a LIZ1200 size standard. LR-PCR products of samples that appeared to be homozygous (only one allele visible) or carriers of alleles of >900 bp (∼250 repeats) were also separated on an 1% agarose gel to exclude or confirm the presence of expanded alleles. Specificity of the PCR products was confirmed by Sanger sequencing of the repeat-flanking region.

### RNA and protein analyses

Primary fibroblasts were established from three affected individuals and four unrelated sex and age matched controls using standard techniques. Cells were cultured as previously described^24^ with RNA extracted (RNeasy Mini Kit, #74104, QIAGEN) and cDNA generated (QuantiTect Reverse Transcription Kit, #205313, QIAGEN) according to the manufacturer’s instructions. Gene expression was analyzed using TaqMan Gene Expression Assays designed to identify all *FGF14* mRNA isoforms (Hs00738588, Thermo Fisher Scientific) or specifically the brain isoform 1b (Hs02888324, Thermo Fisher Scientific) according to the manufacturer’s protocol. Protein was isolated and analyzed by Western blot as previously described^25^ utilizing an antibody directed against FGF14 (UC Davis/NIH NeuroMab, #75-096).

### Statistical analyses

Linear regression was performed in R (version 4.0.5) using the base function [lm()] to determine the correlation between age at onset and RE expansion length. A linear equation was determined from the output of the lm function. Fisher’s exact test was used to test for enrichment of *FGF14* RE >(GAA)_250_ in affected individuals versus controls.

## RESULTS

### Participant recruitment

The workflow for this study is summarized in Figure 1. In the first step, 47 adults with ataxia were recruited and underwent short-read genome sequencing, in addition to subsequent LR-PCR and RP-PCR analysis. All were singletons without a verified family history of ataxia. Second, an additional Australian cohort of 48 individuals was recruited and analyzed by LR-PCR and RP-PCR only. All individuals (n=95) were diagnosed with ataxia based on clinical examination by a neurologist and included 53 female/42 male individuals with adult-onset ataxia (mean AAO 54+/-14 years, range 24-72 years). Individuals were excluded if there was clinical suspicion of an acquired cause of ataxia, based on history of acute injury or illness, toxic exposure, or rapid onset. All samples were screened for pathogenic REs in SCA1, SCA2, SCA3, SCA6 and SCA7 via diagnostic ataxia panel testing prior to research based genetic testing.

**Figure 1.**
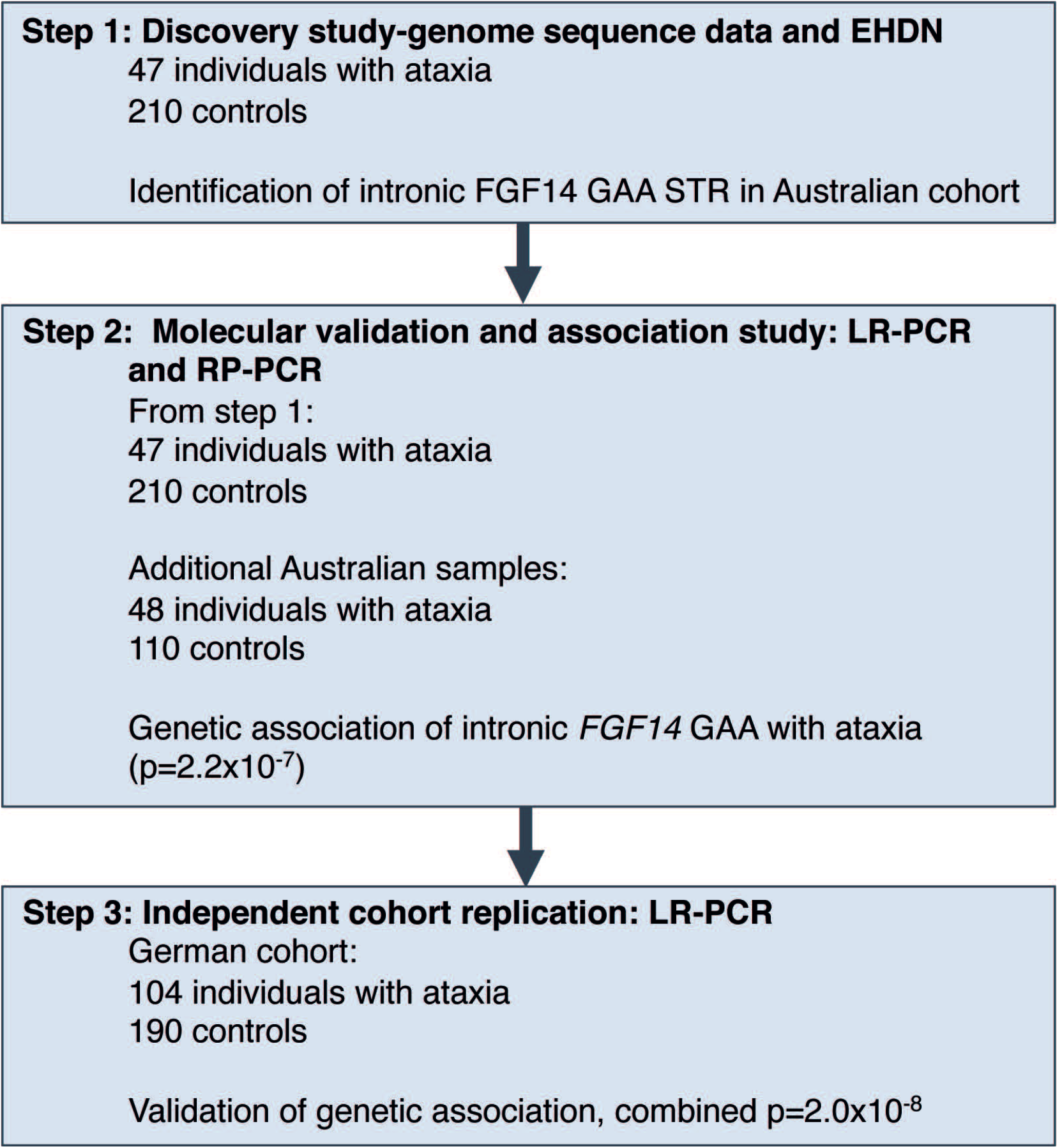
Overview of the study and investigations performed.

### Discovery of a novel repeat expansion in *FGF14*

Given the enrichment of REs as a genetic cause of ataxia, the sequence data of the 47 affected individuals (discovery cohort) was screened for novel REs using ExpansionHunter Denovo (EHDN), compared to 210 unrelated controls. This analysis identified enrichment of an expanded GAA repeat in intron 1 of *FGF14* in participants versus controls, with an outlier z-score of 21.6. Point mutations resulting in haploinsufficiency for *FGF14* are a known rare cause of autosomal dominant spinocerebellar ataxia 27 (SCA27)^26^, however REs in this gene have not been previously identified. The (GAA)_n_ STR is located in the hg38 reference genome at chr13:102161577-102161726, and is reported to comprise 50 pure GAA motifs. Plotting anchored in-repeat reads (a-IRR) from EHDN shows that the STR is present in affected individuals and controls, with seven outliers from the affecteds (a-IRR > 30, Figure 2A) compared to 210 controls (Figure 2B). Large a-IRRs are correlated with longer expansion.^9^ EHDN detected the STR in 21 of 47 affected individuals (44.7%) and 67 of 210 controls (31.9%). The individuals in which no a-IRR was reported either do not have the *FGF14* STR, or it is <150bp (50 repeats) in length as this is the minimum size detectable by EHDN (see below). Visualization of the genome sequence reads in IGV confirmed the likely presence of an expanded (GAA)_n_ STR in intron 1 of *FGF14* (Figure S1). To further characterize the locus prior to proceeding to molecular validation, genome sequence data was analyzed with exSTRa and ExpansionHunter (Figure S2). Results from exSTRa (Figure S2A) indicate that a (GAA)_n_ STR exists at the locus in a range of sizes in the general population, with many controls displaying a high proportion of reads containing full 150bp-reads of GAA repeats. As a result, outlier analysis using exSTRa is not feasible for this locus. In addition, genotyping with ExpansionHunter identifies (GAA)_n_ REs in the 100 repeat range in both affected individuals and controls, although density plots indicate a shift in the distribution towards larger repeats in the affected individuals (Figure S2B).

**Figure 2.**
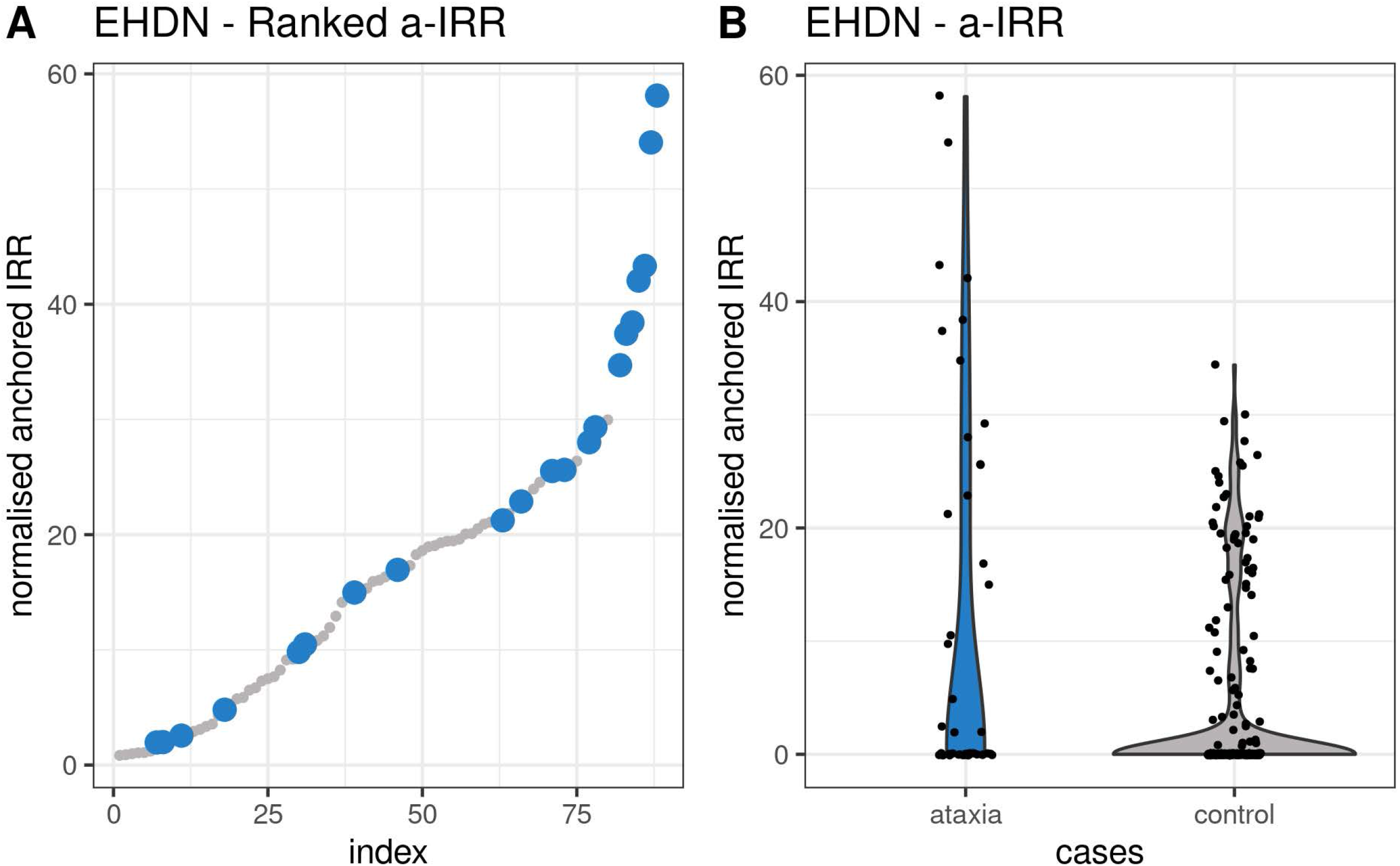
RE analysis with EHDN identifies a potential novel intronic GAA RE in *FGF14* enriched in the ataxia cohort. (A) Summary of ranked anchored-IRR findings for affected individuals (blue) and controls (grey) for the intronic GAA STR in *FGF14*. (B) Violin plot showing the difference in distribution of the anchored-IRR for affected individuals (blue) and controls (grey) for all individuals with an *FGF14* repeat motif of at least (GAA)_50_. Anchored-IRR for individuals with a repeat less than (GAA)_50_ are not detected by EHDN, hence not all individuals analyzed are represented in these plots.

### Molecular validation of the *FGF14* GAA RE

To confirm expansion of the (GAA)_n_ STR we performed standard PCR and agarose gel electrophoresis of two affected individuals identified by EHDN and two controls predicted to not be expanded. This analysis demonstrated additional products >1000bp in the affected individuals, whereas the controls only showed products less than 300bp (Figure 3A). The predicted PCR product size based on hg38 is 315bp, which includes (GAA)_50_. We then developed locus-specific LR-PCR and RP-PCR assays to investigate the size and motif composition of the *FGF14* STR by capillary array analysis. Consistent with the standard PCR assay, LR-PCR analysis identified a heterozygous expanded allele in both affected individuals, with estimated larger allele sizes of (GAA)_289_ and (GAA)_349_ respectively (Figure 3B). In comparison, estimated allele sizes of (GAA)_22_ and (GAA)_12_ were observed in the controls, matching the standard PCR results. Similarly, we observed an extended saw-toothed ‘ladder’ in affected individuals when the RP-PCR products were analyzed by capillary array, which was absent in controls (Figure 3C). These results suggest that EHDN had correctly identified a novel heterozygous (GAA)_n_ RE in intron 1 of *FGF14* and validated molecular tools were applied to perform genetic characterization in the larger cohort.

**Figure 3.**
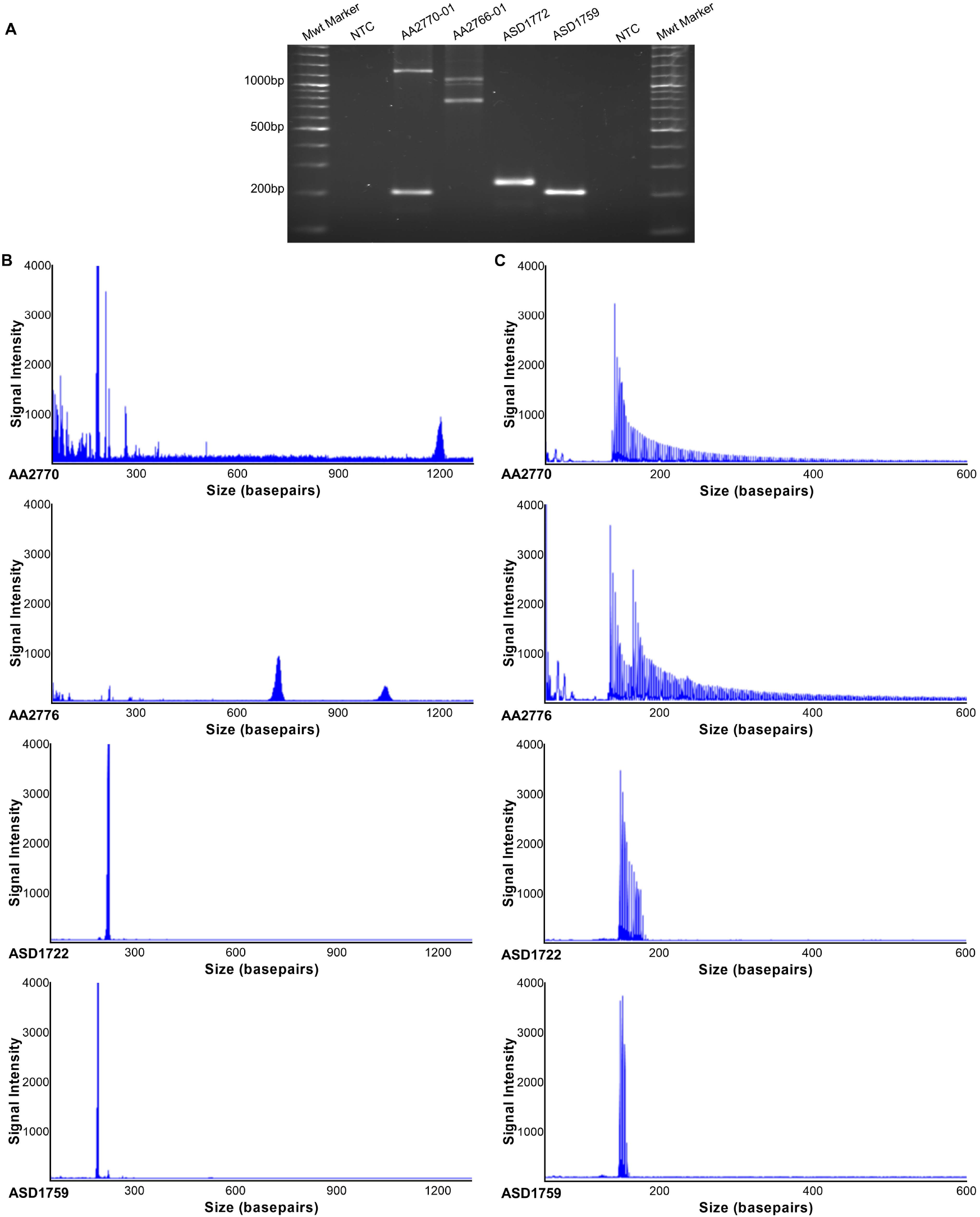
Molecular validation confirms a heterozygous (GAA) RE in *FGF14*. (A) Gel electrophoresis of PCR products spanning the *FGF14* (GAA) STR locus. Affected individuals (AA2770) and AA2766) produced large PCR products of between ∼1000bp and ∼1200bp compared to controls (ASD1722 and ASD1759 <∼300bp). No product was observed in the no-template negative control (NTC). (B) Accurate quantification of (GAA) repeat size was determined by fragment analysis. Individuals with large GAA expansions generated low signal intensity peaks, with staggered peak formations in 3bp intervals >∼1000bp, indicating an allele with a high number of repeats. Electropherograms of non-expanded LR-PCR products displayed a single high intensity spike at <300bp. (C) RP-PCR for the (GAA) RE demonstrated an extended saw-toothed product with three base pair repeat unit size in affected individuals that was truncated or absent in control individuals.

We then performed LR-PCR analysis of all 95 affected individuals and 311 controls, which included an additional 48 affected individuals with no genome sequence data. In total, we identified 13 affected individuals with an allele greater than (GAA)_250_, four of which were (GAA)_>335_ (Figure 4A, Table S3). Analysis of the available genome data (eight individuals) with EHDN confirmed the presence of large *FGF14* (GAA)_n_ expansions. In addition, we screened for pathogenic RE known to cause ataxia using ExpansionHunter and exSTRa, and for pathogenic sequence and copy number mutations in known ataxia genes. No candidate pathologenic variants were identified in these eight individuals. LR-PCR demonstrated only five controls had an allele greater than (GAA)_250_, with the maximum size being (GAA)_300_.

**Figure 4.**
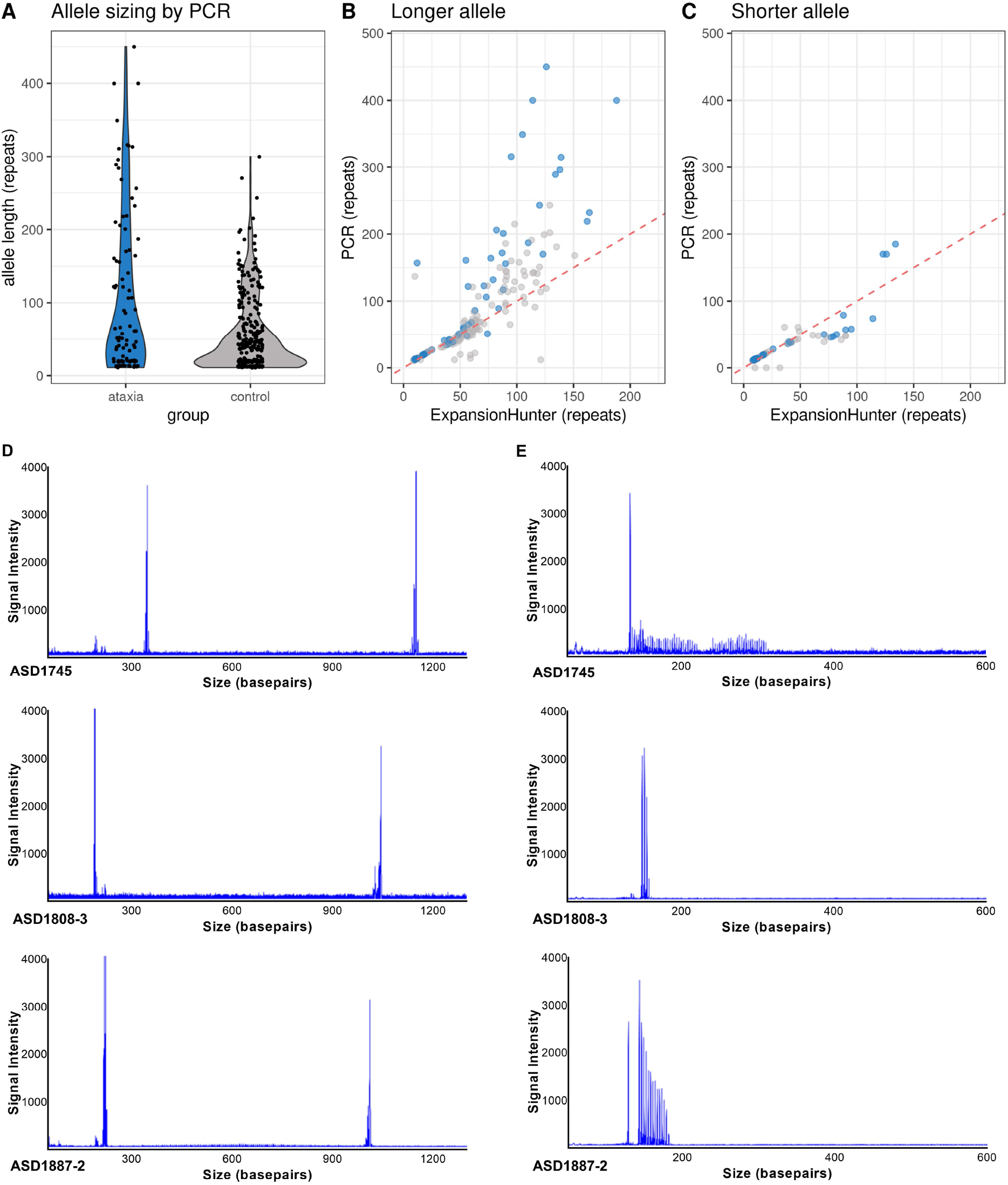
Characterisation of *FGF14* (GAA) RE in study participants. (A) The allele size of the *FGF14* (GAA)_n_ STR was determined in all available affected individuals (n=95) and controls (n=311) using fragment analysis of LR-PCR products. Allele size frequency is indicated on the violin plots, with greater density represented by a wider distribution. Thirteen affected individuals demonstrated pure (GAA)_>250_, compared to only two controls. Comparison of *FGF14* (GAA) repeat length as determined by LR-PCR and ExpansionHunter for the longer (B) and shorter (C) allele demonstrated ExpansionHunter consistently under-estimates repeat size in comparison to molecular sizing [affected individuals (clear blue) and controls (clear grey)]. The dotted red line indicates the y=x relationship representing equivalent sizing. Areas of darker blue indicate overlapping points. (D) Electropherograms from three controls with (GAA)_>250_ display an unusual pattern, with a single high intensity peak observed corresponding to the large allele. (E) RP-PCR analysis demonstrated inconsistent and stunted peak patterns, as opposed to the expected sawtooth ‘ladder’. This pattern is consistent with an impure/alternate repeat motif.

Comparison of allele size estimates by ExpansionHunter and LR-PCR revealed bioinformatic sizing of the RE using short-read GS data was significantly underestimated when the *FGF14* RE was greater than ∼(GAA)_100_ (Figure 4B, C). Interestingly, LR-PCR and RP-PCR revealed a consistent symmetrical peak distribution for all 13 affected individuals with an allele greater than (GAA)_250_ (Figure 3B, C), suggestive of a pure (GAA)_n_ RE. This was confirmed by long-read sequencing (Figure S3), with expanded reads in all five samples analyzed with an RE>(GAA)_250_, being pure GAA. In addition, this analysis also confirmed the RE sizes observed by LR-PCR, with estimates for the two methods differing by less than 5%. In comparison, three of five controls with estimated RE sizes of (GAA)_332_, (GAA)_296_ and (GAA)_282_ displayed unusual LR-PCR peaks (Figure 4D) and RP-PCR traces (Figure 4E). Inspection of the short-read sequence data for these controls clearly demonstrated that the RE was not made up of pure GAA motifs, instead it appeared to consist of GAAGGA hexamer repeats (Figure S4). It is well established that interrupted and impure RE can modify disease penetrance and severity in other autosomal dominant SCA^27-29^ and our analysis raises this possibility that this may apply to disease mediated by GAA expansion in *FGF14*. Considering only pure expanded (GAA)_n_ STRs, we identified 13 affected individuals (13.7%) with an allele greater than (GAA)_250_, but only two controls (0.6%), with pure repeats of (GAA)_272_ and (GAA)_300_, respectively (Figure 4A). Collectively, this data suggests that heterozygous expansion of a pure GAA repeat within intron 1 of *FGF14*, beyond a threshold of ∼250 motifs, represents a common cause of adult-onset ataxia [Fisher’s Exact Test P-value 2.2×10^−7^, Odds Ratio=24.3 (95% CI=5.3-224.7)]. Pure alleles (GAA)_>300_ were exclusively found among affected individuals (n=8) in the Australian cohort.

To test the frequency of the (GAA)_n_ RE in additional populations, we screened a German cohort of 104 individuals with adult-onset ataxia using LR-PCR (Table S3). This analysis identified 15 affected individuals (14.4%) with an allele greater than (GAA)_250_, compared to 10/190 (5.3%) observed in controls [Fisher’s Exact Test P-value 0.014, Odds Ratio=3.03 (95% CI=1.3-7.02)]. Given the differences in the frequency of (GAA)_>300_ in Australian (0/311) and German (7/190) controls, we initially analyzed the combined dataset with stringent criteria, requiring the pathogenic cutoff to be greater than the largest observed control allele [(GAA)_332_]. The results suggest that expanded alleles greater than (GAA)_332_ appear to be pathogenic and fully penetrant [P-value 6.0×10^−8^, OR 72 (95% CI=4.3-1227)]. Similarly, analyzing the combined dataset using the lower size threshold established in the Australian cohort suggests that alleles in the range of (GAA)_250-334_ are likely to be pathogenic [P-value 0.0015, OR 3.6 (95% CI=1.6-7.9)], albeit with reduced penetrance. Large repeats in other SCA are known to be unstable and demonstrate intergenerational variability. For one index participant DNA samples were also available from clinically unaffected parents, allowing evaluation of germline repeat instability. We observed no change in the size of the paternal (GAA)_25_ allele, whereas the maternal allele increased from (GAA)_282_ to (GAA)_315_ (Figure S5). This observation provides anecdotal evidence of meiotic instability of expanded alleles and incomplete penetrance of (GAA)_>250_.

### Profiling of FGF14-GAA in 1000 Genomes Project Data

Next, we applied EHDN to data from the 1000 Genomes Projects to further characterize the composition of the *FGF14* STR locus in diverse populations. EHDN only analyzes reads that contain a pure motif and that are >150bp, therefore any STRs that are shorter than the read length (i.e. <150bp/50 triplet repeats) are not detected by EHDN. We found that the *FGF14* STR >(GAA)_50_ is present in all five super populations represented in the 1000 Genomes project with frequencies of: African (8%), ad-mixed American (22.4%), East Asian (8.5%), South Asian (24.1%) and European (32.3%) (Figure S6A). Furthermore, European and South Asian populations have higher a-IRR on average, while African populations have the lowest counts of a-IRR (Figure S6B), indicating that longer forms of the STR are present across populations, but in different frequencies. In addition, while (GAA)_n_ is by far the most common, EHDN also identified population-specific alternative motifs that are variations of GAA, including GAAGGA and GAAGAAGAAGAAGCA, both of which are present in East Asian and ad-mixed American populations (Figure S6C).

### Analysis of *FGF14* expression in primary fibroblast lines

Two predominant mRNA isoforms are produced by *FGF14*, which result from usage of different first exons. The primary site of expression of the longer isoform (1b) is the brain, with high levels observed in the cerebellum and cortex.^30^ The (GAA)_n_ RE is located within intron 1 of isoform 1b (Figure 5). Therefore, it is possible that the *FGF14* RE can interfere/reduce gene transcription, analogous to how expansion of the only other known (GAA) RE, located in intron 1 of *FXN*, reduces *FXN* expression and causes Friedreich ataxia.^31^ Reduced expression of the mutant *FGF14* allele could lead to haploinsufficiency, the disease mechanism underlying SCA27, caused by heterozygous mutations in *FGF14*.^26^ To test this, we generated primary fibroblasts from three affected individuals with (GAA)_244_, (GAA)_289_ and (GAA)_348_. Expression of both isoforms 1a and 1b is reported to be very low in fibroblast cell lines (GTEx) and indeed we were unable to reliably detect either isoform using TaqMan Gene Expression Assays and standard qRT-PCR or ddPCR, the latter being an extremely sensitive tool for absolute quantitation of gene expression.^32^ Similarly, Western blot analysis failed to identify FGF14 in protein extracts from participant or control lines, reflecting the low gene expression (data not shown).

**Figure 5.**
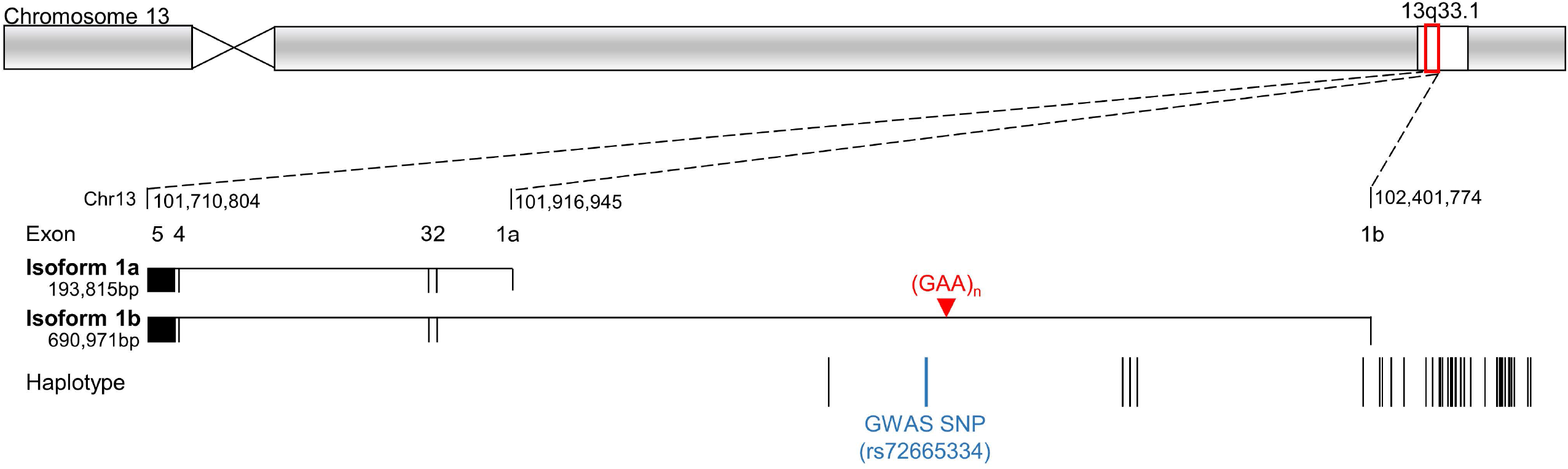
*FGF14* genomic region, haplotype and isoform structure. Schematic representation of the chromosomal location of *FGF14* to 13q33.1, illustrating the expression sites of two mRNA isoforms, 1a and 1b. Exon one of isoform 1b is located 5’ of 1a, and the (GAA)_n_ STR is located within the intervening intron. The remaining exons 2-5, are common to both isoforms. The SNP (rs72665334) identified by GWAS to be associated with down beat nystagmus is located ∼10kb 3’ of the (GAA)_n_ STR. A common ancestral haplotype was determined for three Australian cases with (GAA)_>250_ by comparing variant sharing around the *FGF14* STR (gnomAD AF<0.1) (specific haplotype details in Table S3).

### Clinical findings

A pure *FGF14* RE >(GAA)_250_ was identified in 13 Australian individuals with cerebellar ataxia (CA). The sex distribution was 5 female to 8 male and mean age at symptom onset was 61 years (range 46-77 years). Participants were evaluated at a mean age of 69 years (range 54-81), after a mean period of 8.5 years (range 1-16) since symptom onset. All displayed a range of clinical ataxic phenomena, with MRI scanning revealing cerebellar atrophy in five of the 13 (Table 1). Six of 10 assessed had evidence of vestibular hypofunction on video Head Impulse Test (vHIT)^33^, five were bilaterally hypoactive and one was unilaterally hypoactive. An abnormal video Visually Enhanced Vestibulo-Ocular Reflex (VVOR) was seen in those with bilateral vestibular hypofunction.^34^ Where formally assessed with a pure tone audiogram, four of 10 had hearing loss consistent with presbycusis, four of 10 with noise-induced hearing loss (HL), one had unilateral post-traumatic HL and one had early-onset HL (the details of which were unavailable). Additional extracerebellar features are described in Table 1.

**Table 1.**

| ID | (GAA). size | Demographics |  |  |  | Cerebellar clinical features |  |  |  |  |  | Vestibular function | Neurophysiology |  | MRI | Extracerebellar features | Core phenotype |
| --- | --- | --- | --- | --- | --- | --- | --- | --- | --- | --- | --- | --- | --- | --- | --- | --- | --- |
|  |  | Sex | Age at onset (decade) | Age at assessment (decade) | Disease duration (decade) | Upper limb ataxia | Lower limb ataxia | Gait ataxia | Oculomotor abnormalities | Truncal ataxia | Cerebellar dysarthria | Hypofunction on vHIT | Sensory +/- or motor impairment on NCS | Autonomic nervous system dysfunction | Cerebellar, brain stem or other atrophy |  |  |
| AA2807 | 450 | M | 5 | 6 | >1 | ✓ | ✓ | ✓ | ✓ | ✓ | ✓ | - | X | - | X | X | CA^ |
| AA2903 | >400 | F | 6 | 7 | >1 | ✓ | ✓ | ✓ | ✓ | X | ✓ | unilateral | X | ✓ | X | hyper-reflexia | CAUV, ANS and hyper-reflexia |
| AA2831 | >400 | M | - | 7 | - | X | - | ✓ | - | - | - | - | - | - | anterior cerebellar atrophy | spasticity | CA* |
| AA2770 | 348 | F | 6 | 8 | >1 | ✓ | ✓ | ✓ | ✓ | X | ✓ | X | X | - | superior and posterior vermal, mild hemispheric atrophy | chronic cough | CA and cough |
| AA0441 | 317 | M | 7 | 8 | <1 | ✓ | ✓ | ✓ | ✓ | X | ✓ | X | X | - | X | X | CA |
| AA2895 | 312 | M | 5 | 6 | <1 | X | ✓ | ✓ | ✓ | X | - | - | - | - | generalised cerebellar atrophy, principally vermis | lower limb hyper-reflexia, spasticity | CA^ and spasticity |
| AA2775 | 310 | M | 6 | 7 | >1 | ✓ | ✓ | ✓ | ✓ | X | ✓ | bilateral | X | ✓ | anterior and posterior vermal, mild hemispheric atrophy | lower limb hyper-reflexia | CABV, ANS and hyper-reflexia |
| AA2809 | 309 | M | 8 | 8 | <1 | ✓ | X | ✓ | ✓ | X | X | bilateral | X | ✓# | X | lower limb hyper-reflexia, parkinsonism | CABV, hyper-reflexia and parkinsonism |
| AA2845 | 298 | F | 7 | 8 | <1 | X | X | ✓ | ✓ | X | X | X | X | - | X | X | CA |
| AA2766 | 289 | M | 7 | 8 | >1 | ✓ | ✓ | ✓ | ✓ | X | ✓ | bilateral | X | ✓ | superior and posterior vermal, mild hemispheric atrophy | chronic cough | CABV, ANS and cough |
| AA2908 | 281 | F | 6 | 8 | >1 | ✓ | ✓ | ✓ | ✓ | ✓ | ✓ | bilateral | - | - | X | clinical ANS dysfunction | CABV and clinical ANS |
| AA2933 | 280 | F | 8 | 9 | <1 | X | X | ✓ | ✓ | X | X | X | - | - | X | X | CA |
| AA2926 | 270 | M | 7 | 8 | <1 | ✓ | ✓ | ✓ | ✓ | X | X | bilateral | - | - | X | X | CABV |
| L-18362 | ~460 | M | - | 8 | - | ✓ | ✓ | ✓ | ✓ | ✓ | ✓ | bilateral | X | - | cerebellar atrophy, primarily, superior vermis | spasticity | CABV and spasticity |
| L-17665 | ~430 | F | 5 | 6 | <1 | X | X | X | ✓ | X | X | bilateral | X | - | X | X | CABV |
| L-17672 | ~430 | M | 6 | 6 | <1 | X | ✓ | ✓ | ✓ | X | X | X | X | - | X | X | CA |
| L-18384 | ~430 | M | 7 | 7 | <1 | X | X | ✓ | ✓ | X | X | bilateral | X | - | - | X | CABV |
| L-20363 | ~400 | M | 7 | 9 | >3 | ✓ | ✓ | ✓ | ✓ | - | X | unilateral | X | - | - | X | CAUV |
| L-15166 | ~350 | M | 5 | 5 | <1 | ✓ | ✓ | ✓ | ✓ | - | ✓ | - | - | - | frontal atrophy | spasticity | CA^, spasticity |
| L-15764 | ~350 | M | 7 | 8 | <1 | ✓ | ✓ | ✓ | ✓ | ✓ | ✓ | bilateral | X | - | moderate global atrophy, predominantly cerebellar | X | CABV |
| L-14630 | 342 | M | 6 | 8 | >1 | X | X | ✓ | ✓ | X | X | - | - | - | X | clinical ANS dysfunction | CA^ and clinical ANS |
| L-10410 | 315 | F | - | - | - | - | - | ✓ | - | - | - | - | - | - | - | - | CA |
| L-15891 | 312 | M | 8 | 8 | <1 | ✓ | ✓ | ✓ | X | - | X | bilateral | - | - | X | clinical ANS dysfunction | CABV and clinical ANS |
| L-15754 | ~290 | F | 8 | 9 | >1 | ✓ | ✓ | ✓ | ✓ | - | X | bilateral | - | - | X | clinical ANS dysfunction | CABV and clinical ANS |
| L-15713 | ~270 | M | 8 | 8 | <1 | ✓ | ✓ | ✓ | ✓ | ✓ | ✓ | bilateral | X | - | global brain and cerebellar atrophy | clinical ANS dysfunction | CABV and clinical ANS |
| L-15739 | ~270 | F | 8 | 8 | <1 | X | X | ✓ | ✓ | - | X | bilateral | X | - | global brain atrophy | X | CABV |
| L-15629 | ~270 | M | 8 | 9 | <1 | ✓ | ✓ | ✓ | ✓ | ✓ | X | X | - | - | global brain and cerebellar atrophy | X | CA |

vHIT=video head impulse test, NCS=nerve conduction study, ANS=autonomic nervous system, M=male, F=female, ✓=present, X=absent, -=no data available, CA=cerebellar ataxia, CABV=cerebellar ataxia and bilateral vestibulopathy, CAUV=cerebellar ataxia and unilateral vestibulopathy, ^= vHIT not performed, \*=cerebellar ataxia no other phenotypic information available, ✓#=present on anti-hypertensive medication. Note clinical details such as age at onset have been presented in a form consistent with the requirements of the publisher regarding potentially identifying information.

### Neurophysiology

Both motor and sensory studies were normal in the six affected individuals in whom a detailed nerve conduction study was performed, apart from two who had focal median motor and sensory slowing across the wrists, indicating asymptomatic median neuropathies at the wrists (i.e. asymptomatic carpal tunnel syndrome). The findings were normal in the individual who had only lower limb studies. The findings in the one individual who had lower limb studies and limited upper limb studies were normal apart from an asymptomatic median neuropathy at the wrist. Small nerve fibre studies were normal in the six affected individuals in whom they were performed and provided no evidence of a somatic small fibre neuropathy. All four individuals who underwent tilt table testing had borderline to mild asymptomatic orthostatic falls of the systolic blood pressure of 18-37 mmHg. One of these individuals who also had a reduced heart rate response to deep breathing and standing, was taking anti-hypertensive medication at the time of testing and given this potential confound, we can only conclude that three displayed evidence of orthostatic hypotension. In addition, one participant had a reduced heart rate response to deep breathing and standing. Overall, the neurophysiology investigations failed to show consistent evidence of somatic large or small fibre neuropathy or neuronopathy. The tilt table and autonomic testing suggests likely involvement of the autonomic nervous system.

### Age At Onset analysis

RE expansion length is often inversely correlated with disease age at onset. We applied a linear regression model to determine the relationship between *FGF14* RE length and ataxia onset age for the individual cohorts and also for the pooled data. This analysis identified an inverse correlation (R^2^=0.44, p=0.00045, slope =-0.12). The regression suggests that for every (GAA)_10_ increase above 250 repeats, the age of onset is reduced by ∼1.16 year. Similar results were observed when the Australian (R^2^=0.36, p=0.038, slope=-0.10) and German (R^2^=0.55, p=0.0058, slope=-0.14) cohorts were analyzed independently (raw data not shown due to publisher restrictions).

## DISCUSSION

Cerebellar ataxia (CA) may be defined as a disturbance of the normal co-ordination of movements and comes about when there is an impairment of cerebellar function. CA have multiple etiologies and many are associated with progressive and debilitating illness. Pathogenic repeat expansions collectively represent the most common genetic cause of CA and while they have traditionally proven difficult to both discover and assess diagnostically using standard molecular tools and practices, recent advances in genomic technologies have begun to address these limitations.^35,36^ Here we demonstrate the power of new bioinformatic tools and second and third generation sequencing platforms by identifying and characterizing a novel pathogenic (GAA)_n_ RE within intron 1 of *FGF14* in a significant proportion of unresolved individuals with adult-onset ataxia. We replicate the initial finding made in an Australian cohort in an independent German cohort. Interestingly, seven (GAA)_301-332_ alleles were observed in German controls, compared to none in the Australian samples. This may reflect a difference in population structure or the fact that some of these may represent non-pure alleles, which were identified and removed from the Australian dataset. Irrespective, our data strongly supports a fully penetrant threshold of (GAA)_>335_ for this RE and also supports (GAA)_>250_ as likely pathogenic, albeit with reduced penetrance. However, defining the pathogenic range will require additional, larger studies in aged controls since the disease onset can occur very late in life; onset ages in the eighth decade were observed in our cohorts. Given that de-identified DNA samples for ‘healthy’ controls, such as the HRC1 panel used in this study, are often acquired many years prior to potential disease onset, it is difficult to determine if an expanded allele is non-penetrant, below a ‘pathogenic’ threshold or the individual is yet to manifest the condition. In addition, this study has highlighted the likely need to determine both the size and structure (repeat sequence) of expanded *FGF14* alleles as a part of molecular diagnosis. Achieving a genetic diagnosis confers a number of benefits including definitive diagnosis, improved prognostication, possible implications for reproductive choices and identification of gene therapy targets. We anticipate increasing application of third generation (long-read) sequencing technologies in diagnostic facilities to meet the challenges of accurate genetic diagnosis for *FGF14* and indeed the majority of RE-mediated disorders. We demonstrate that *FGF14* can be efficiently targeted and screened with the adaptive sampling technique afforded by ONT sequencing.

While this study has a cross-sectional cohort design, our results suggest that *FGF14* RE-mediated disease can result from either *de novo* expansion or familial inheritance of an expanded pathogenic allele. Three individuals share a core haplotype spanning ∼395kbp, suggesting distant relatedness and inheritance of a founder RE. Indeed, the haplotype includes rs72665334, a single nucleotide variant identified by GWAS as associated with down beat nystagmus (DBN).^37^ Given that DBN is the most common form of abnormal central nervous system-mediated nystagmus^38^ and is observed in a broad range of CAs, it is not surprising that it is a feature of *FGF14* RE-mediated disease. However, the haplotype result raises the intriguing possibility that DBN-associated rs72665334, present in the global population with an aggregate allele frequency of ∼5% (dbSNP-ALFA), is actually the result of the variant being on a founder haplotype in linkage disequilibrium with a *FGF14* (GAA)_n_ RE. Although this study does not include *FGF14*-linked multiplex families to determine intergenerational RE stability, our analysis of control cohorts and the 1000 Genomes project (Figure 4, S6) has demonstrated that the *FGF14* (GAA)_n_ STR is highly polymorphic in the population. Moreover, we observed intergenerational expansion of a (GAA)_282_ allele in an unaffected female to (GAA)_315_ in her affected daughter (Figure S5).

Members of the FGF family possess broad mitogenic and cell survival activities, and are involved in a variety of biological processes, including embryonic development, morphogenesis, tissue repair, tumor growth and invasion. *FGF14*, and in particular isoform 1b, is widely expressed in the developing and adult brain, with highest levels observed in the cerebellum and cerebral cortex.^30^ The protein binds to voltage-gated Na+ (NaV) channels and promotes their localization to the proximal region of the axon, providing the fine-tuned regulation necessary for normal functioning.^39,40^ The development of ataxia and paroxysmal dyskinesia in mice lacking *Fgf14* was the first direct evidence potentially linking the gene to human disease.^41^ Notably, FGF14 is required for spontaneous and evoked firing in cerebellar Purkinje neurons and for motor coordination and balance.^42,43^ Therefore, although our functional studies of primary fibroblasts were inconclusive, we hypothesize that the pathogenic mechanism underlying *FGF14* (GAA)_n_-mediated ataxia is reduced functional protein due to decreased expression of the allele with the RE. Such a mechanism is somewhat analogous to Friedreich ataxia, the only other known example of an ataxia mediated by a (GAA)_n_ RE. This mechanism is also consistent with our observation that GAAGGA repeats, such as (GAAGGA)_166_, equivalent in size to (GAA)_332_, are not associated with ataxia (Figure 3). GGA interruptions in long GAA repeats have been shown to inhibit the formation of triplex and sticky DNA structures and alleviate transcription inhibition.^44^

Our proposed genotype-phenotype correlation is consistent with the observation that point mutations and other non-RE genetic variants in *FGF14* cause autosomal dominant cerebellar ataxia (SCA27, also known as ATX-FGF14).^26,45^ The underlying pathogenic mechanism in SCA27 is a 50% reduction in the normal level of functional protein i.e. haploinsufficiency. SCA27 is a rare form of inherited CA and is described in less than 30 families.^46^ Affected individuals tend to present with early onset CA associated with widespread tremors, dyskinesias, significant cognitive impairment and psychiatric features^26,47,48^, the latter potentially severe.^49^ Less commonly reported signs are microcephaly, clinodactyly and pes cavus.^45^ In contrast, people with *FGF14* (GAA)_n_-mediated ataxia present at a later age with a less severe disease course, outcomes predicted by a reduction but not complete loss of expression of the mutant *FGF14* allele. A similar genotype-phenotype correlation occurs with *CACNA1A*: heterozygous point mutations are associated with episodic ataxia, which almost exclusively manifests prior to 20 years of age whereas a (CAG)_n_ RE in the gene is associated with an adult-onset, slowly progressive spinocerebellar ataxia (SCA6).^50^

The core phenotypes associated with *FGF14* (GAA)_n_-mediated ataxia are pure CA and cerebellar ataxia with bilateral vestibulopathy (CABV), with the variable presence of other features including hyper-reflexia and autonomic dysfunction. Notably, both CA and CABV are conspicuously under-represented in individuals with ataxia in whom a genetic diagnosis is achieved. For example, they are rarely observed in *RFC1*-mediated disease^51,52^, which is thought to be the most common inherited cause of CA.^53^ The identification and high prevalence of *FGF14* (GAA)_n_-mediated ataxia, a novel RE mutation mechanism in a gene previously known to cause a related phenotype, may account for a significant proportion of unresolved individuals with ataxia and fill this diagnostic vacuum. A noteworthy feature of our study is that several participants partially fulfilled criteria for CANVAS with cerebellar ataxia and vestibulopathy, some with additional cough and autonomic failure but all lacking large fibre sensory neuropathy. Our identification of a *FGF14* (GAA)_n_ RE in these patients provides an alternative genetic cause in patients with CANVAS-like syndromes negative for biallelic RFC1 RE. A feature of other RE-mediated CAs such as SCA1,2,3,6,7^54^ and Friedreich Ataxia^55^ is an inverse relationship between repeat length and age at disease onset. Identifying such a relationship can aid prognostication, for example in SCA3 larger expansions are also associated with a more rapid disease progression and varying phenotypes.^6^ Despite the relatively small cohort sizes, we identified a negative correlation between age at onset and repeat length (R^2^=0.44, p=0.00045, slope = -0.12, raw data not shown due to publisher restrictions).

In conclusion, we show that a heterozygous (GAA)_n_ RE located in intron 1 of *FGF14* is a common cause of adult-onset ataxia, in particular individuals with pure CA or CABV phenotypes, a cohort in whom genetic diagnosis is relatively rarely forthcoming. Consistent with the nomenclature guidelines widely utilized by clinicians, we propose the name SCA50 for this newly described disorder. In accordance with the guidelines established by the Movement Disorder Society Task Force for genetic movement disorders ^56,57^ the nomenclature for this ataxia is ATX-FGF14. Our data suggests that (GAA)_>250_ be considered as pathogenic, with acknowledgement of potential incomplete penetrance. It is not currently possible to define an upper size whereby expansions might be classified as fully penetrant, but our analysis of >500 controls suggests (GAA)_>335_. Generating more specific pathogenic ranges will require analysis of large cohorts and families. In addition, given both size and composition appear to be key mediators of pathogenicity, fully resolving these key questions will require application of advanced sequencing technologies, in addition to more traditional molecular tools such as LR-PCR and RP-PCR. Notably, despite the advances in RE diagnostics being enabled by short-read genome sequencing^58^, this study highlights limitations of short-read data and bioinformatic tools such as ExpansionHunter and exSTRa to interrogate the structure of the (GAA)_n_ RE and the difficulty in interrogating larger REs (>150bp). Application of multiple bioinformatic tools, including EHDN, in combination with long-read sequencing technologies will be required to unravel the full genotype-phenotype relationships in SCA50.

## Data Availability

All data produced in the present work are either contained in the manuscript or available upon reasonable request excluding material unable to be shared for ethics/confidentiality purposes

## SUPPLEMENTAL DATA

### CONFLICTS OF INTEREST

The authors declare no conflicts of interest.

## ACKNOWLEDGMENTS

This work was supported by the Australian Government National Health and Medical Research Council grants (GNT2001513 and MRFF2007677) to PJL, MBD and HR. HR was supported by a NHMRC Emerging Leadership 1 grant (1194364) and MB was supported by an NHMRC Leadership 1 grant (1195236). Additional funding was provided by the Independent Research Institute Infrastructure Support Scheme, the Victorian State Government Operational Infrastructure Program and the Murdoch Children’s Research Institute. NB and KL are supported by research grants from the Deutsche Forschungsgemeinschaft (DFG, FOR 2488). We would like to thank Frauke Hinrichs for technical support and Christoph Helmchen, Max Borsche and Meike Kasten for help with the recruitment of the German participants.

## WEB RESOURCES

cavalier: https://github.com/bahlolab/nf-cavalier

dbSNP-ALFA: https://www.ncbi.nlm.nih.gov/snp/docs/gsr/alfa/

exSTRa: https://github.com/bahlolab/exSTRa/blob/master/inst/extdata/repeat_expansion_disorders_hg38.txt

Genotype-Tissue Expression (GTEx) project: https://gtexportal.org/home/

Genome Aggregation Database (gnomAD): http://gnomad.broadinstitute.org/

Integrative Genomics Viewer (IGV): http://software.broadinstitute.org/software/igv/

Online Mendelian Inheritance in Man: http://www.omim.org/

PanelApp Australia: https://panelapp.agha.umccr.org/

PanelApp UK: https://panelapp.genomicsengland.co.uk/

UCSC Genome Bioinformatics database: https://genome.ucsc.edu/

Varsome: https://varsome.com

### ACCESSION NUMBERS

The ClinVar submission for the FGF14 variants reported in this paper is SCV002576522.

---

**Figure S1:**
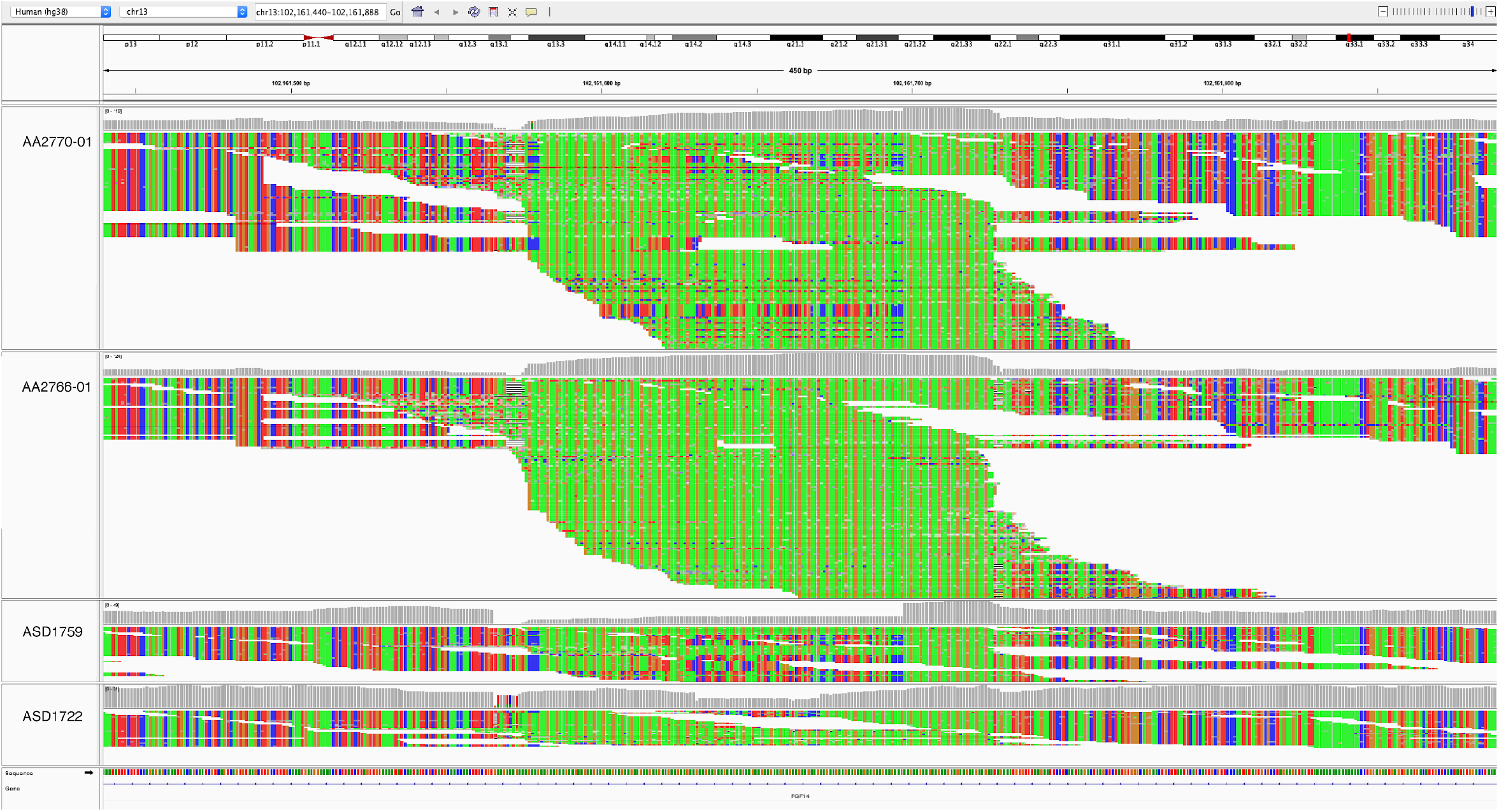
IGV snapshots of the (GAA)_n_ locus in *FGF14*. Visualization of the GS reads in IGV for individuals with an expanded (GAA) RE (top two images) compared to controls (bottom two images).

**Figure S2:**
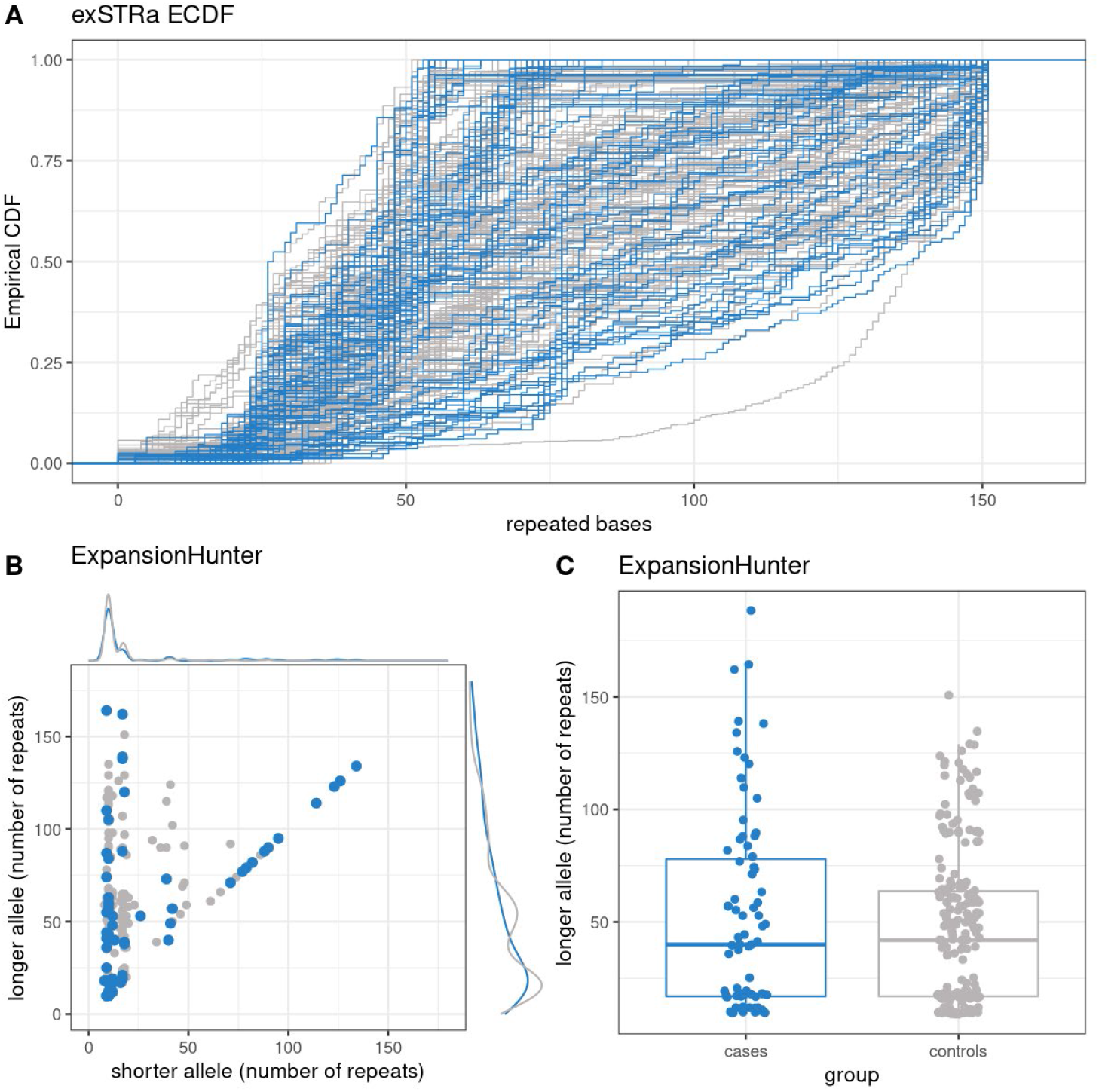
Plots of exSTRa and ExpansionHunter outputs for the *FGF14* (GAA) locus. The *FGF14* (GAA) STR was profiled in cases (blue) and controls (grey) using bioinformatics tools, showing inability of bioinformatic tools to accurately estimate the size of the *FGF14* STR, particularly for alleles with >(GAA)_100_. (A) exSTRa ECDF plot fails to distinguish outliers, due to the high variability of and large size of the STR in the general population. (B) Size estimates of the *FGF14* (GAA) STR were determined using ExpansionHunter. Density plots are show the allele repeat distributions for cases (blue) and controls (grey) for the longer (y-axis) and shorter (x-axis) alleles. (C) Comparison of longer allele size determined by ExpansionHunter in cases versus controls.

**Figure S3:**
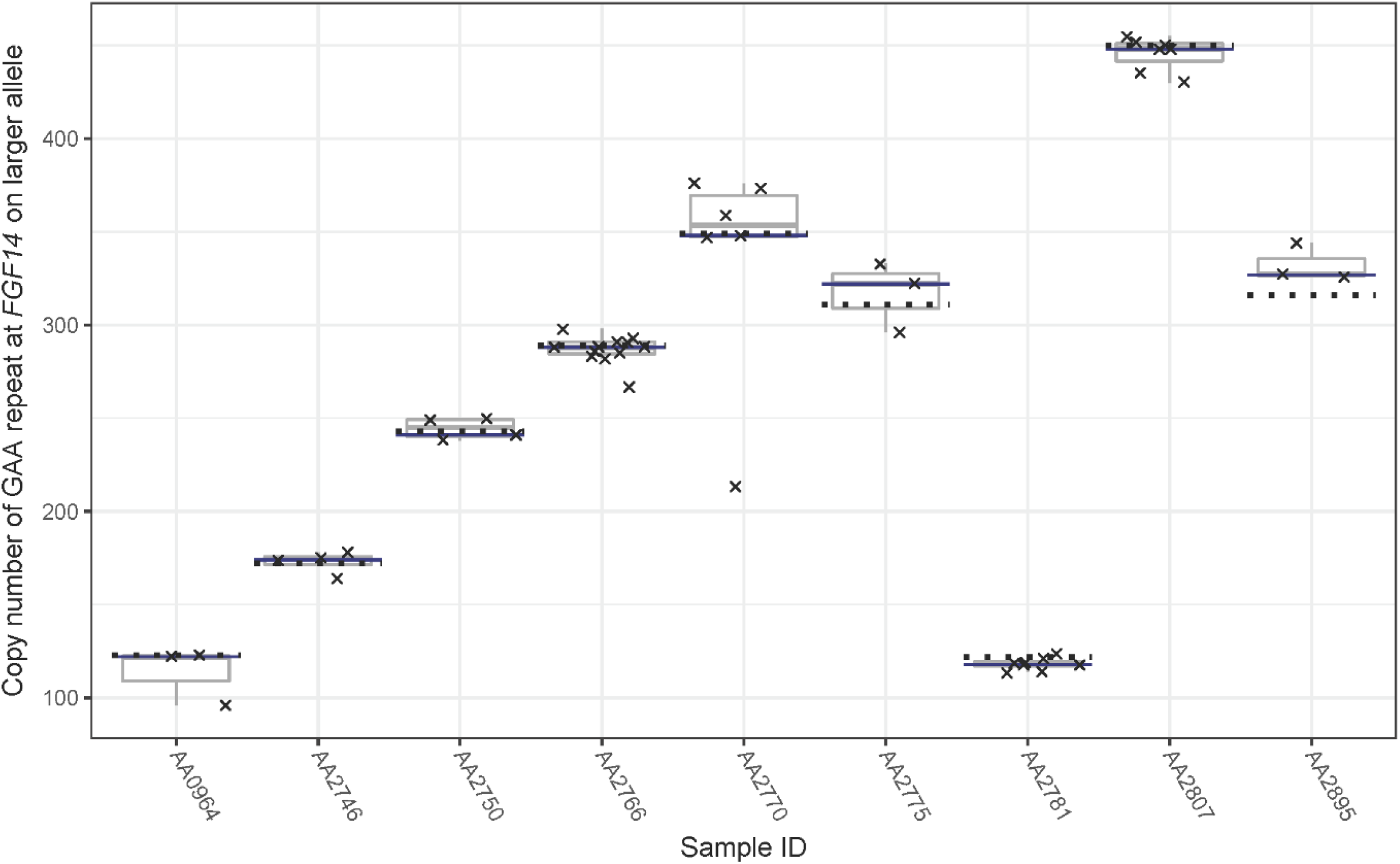
Nanopore repeat size estimates concur with PCR estimates. Eight samples were sequenced with Oxford Nanopore MinION adaptive sequencing targeting *FGF14*. The (GAA)_n_ repeat was genotyped in the long-read sequencing data using tandem-genotypes. Repeat sizes from Nanopore sequencing are shown for each read (crosses) and as a distribution (box plot), along with the tandem-genotypes (blue line) and PCR (dotted line) size estimates for each sample.

**SF4:**
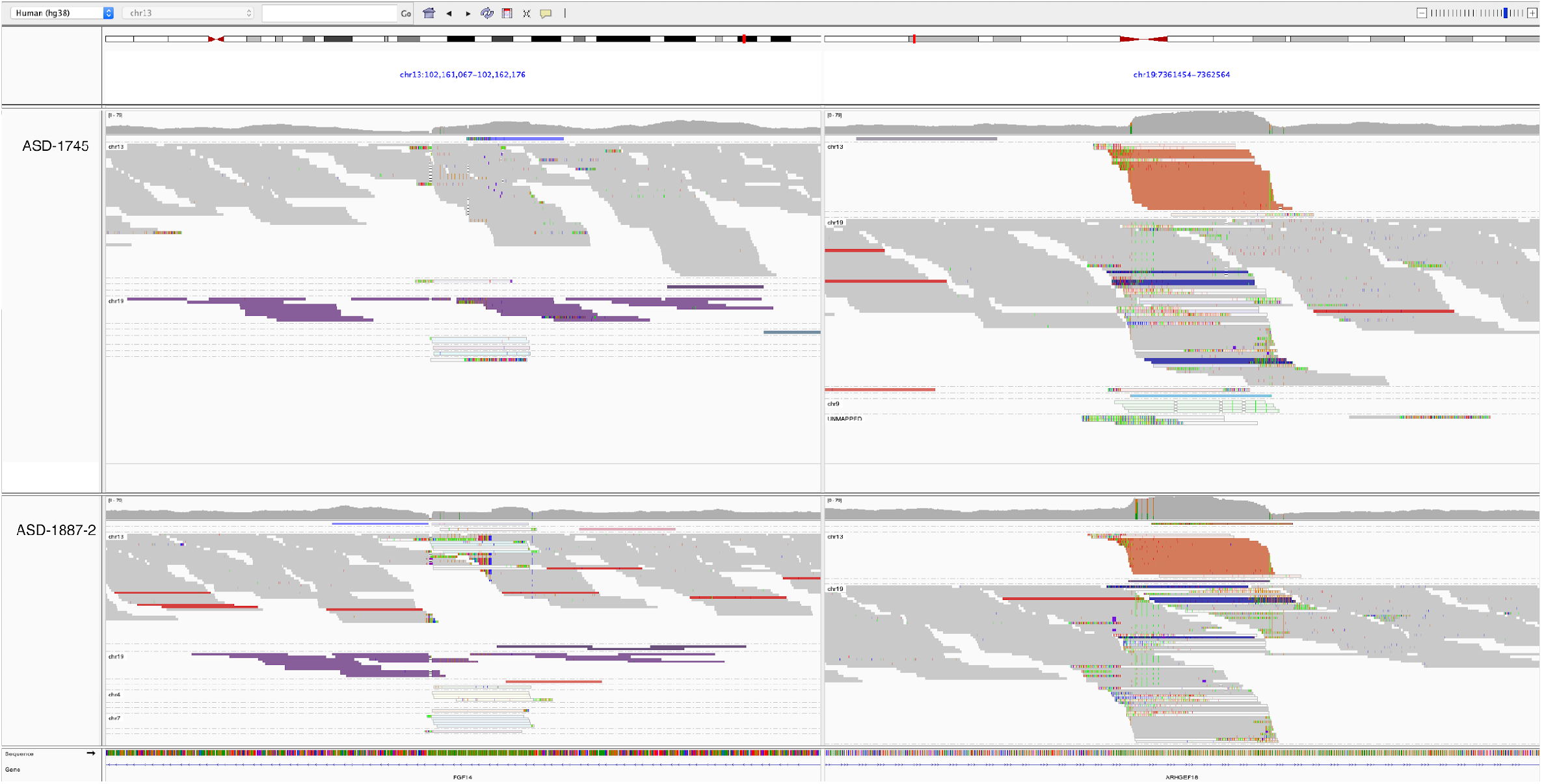
IGV snapshot of controls with unusual RP-PCR patterns. Visualization of the genome sequence reads in IGV for controls with an expanded GAAGGA RE at the *FGF14* (GAA)_n_ STR (left panel). Reads coloured in purple uniquely map to the *FGF14* locus, however their read pairs map to a GAAGGA motif in another region of the genome (right panel, reads in orange).

**Figure S5:**
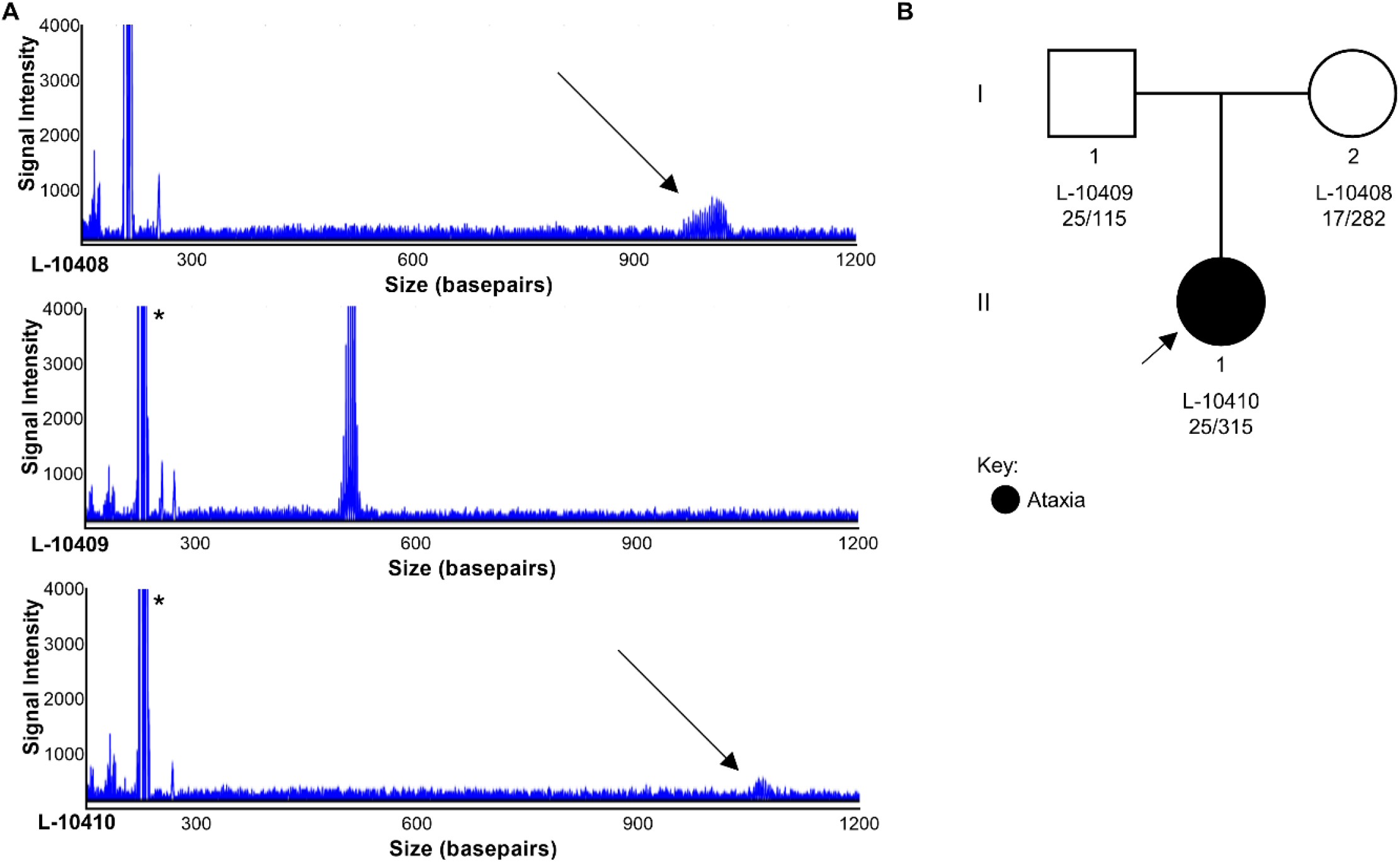
Intergenerational expansion of *FGF14* (GAA)n STR. LR-PCR electropherograms of *FGF14* STR by capillary array (A) for II-1 (L-10410) and unaffected parents (L-10408 and L-10409) (B). Allele sizing for L-10408 at this locus demonstrated that the paternal allele (GAA)_25_ (indicated by *) was inherited unchanged, whereas the maternal allele increased from (GAA)_282_ to (GAA)_315_ in the affected daughter.

**Fig S6:**
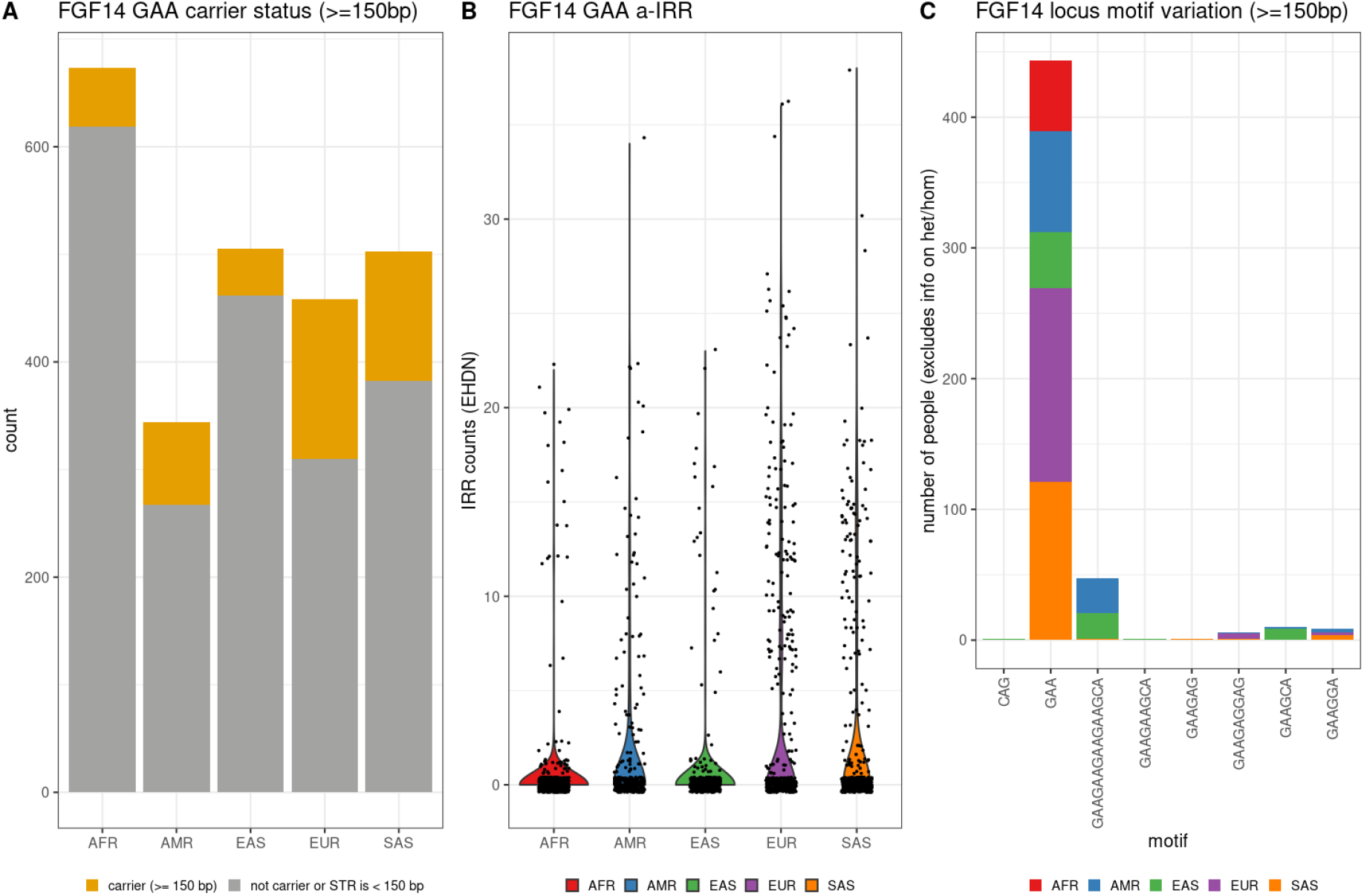
Profiling the *FGF14* (GAA) RE locus in the 1000 Genomes Project. The *FGF14* (GAA) STR locus was profiled in 2483 unrelated individuals from the 1000 Genomes Project from African (AFR), ad mixed American (AMR), East Asian (EAS), European (EUR) and South Asian (SAS) populations. (A) Number of *FGF14* (GAA) STR carriers in the 1000 Genomes Project, by ethnicity. Carriers are defined as having an STR >= 150 bp (or 50 repeats). (B) Violin plot showing the distribution of the a-IRR for the *FGF14* (GAA) STR by ethnicity. Individuals without a reported a-IRR, either because they do not carry the STR, or they carry the STR with <150 bp (or 50 repeats), are coded as zero. (C) Bar chart showing the alternative motifs also detected at the *FGF14* (GAA) locus, split by ethnicity. Only motifs present at >= 150bp in at least one individual are reported.

**Table S1:**
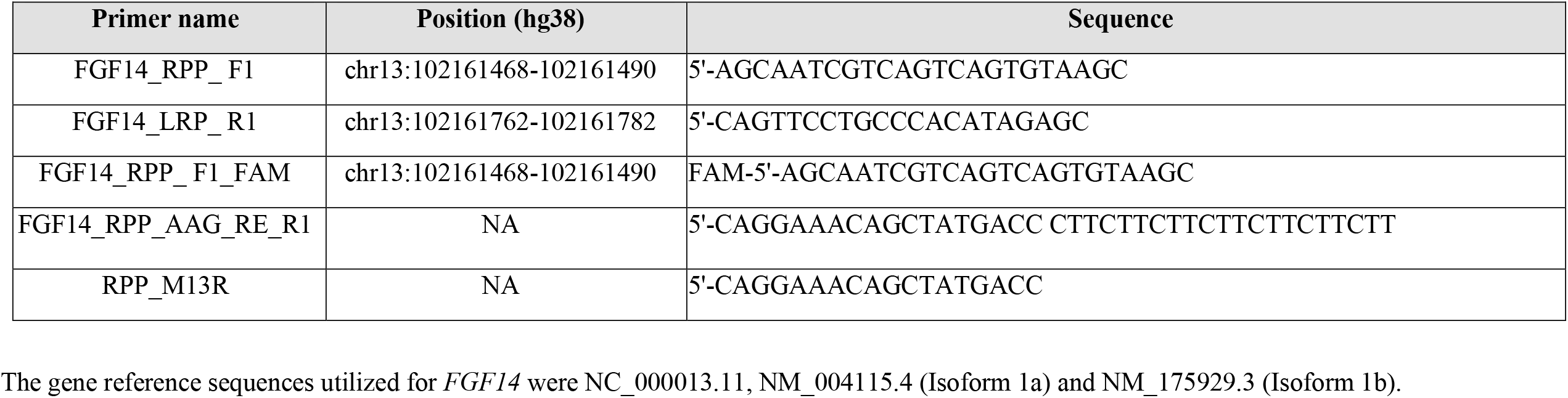
Primer sequences for analysis of *FGF14*.

**Table S2.** *FGF14* allele sizing for individuals with (GAA)_>250_ in Australian and German cohorts.

| Australian Cohort |  |  |  |  |  | German Validation Cohort |  |  |  |  |  |
| --- | --- | --- | --- | --- | --- | --- | --- | --- | --- | --- | --- |
| Affected Individuals |  |  | Controls |  |  | Affected Individuals |  |  | Controls |  |  |
| Alias | Allele |  | Alias | Allele |  | Alias | Allele |  | Alias | Allele |  |
|  | 1 | 2 |  | 1 | 2 |  | 1 | 2 |  | 1 | 2 |
| AA2807 | 170 | 450 | ASD1745 | 65 | 332* | L-18362 | 28 | 460 | L-3657 | 49 | 332 |
| AA2831 | 74 | 400 | C0063 | 12 | 300 | L-14575 | 29 | 460 | L-3656 | 315 | 330 |
| AA2903 | 19 | 400 | ASD1808 | 12 | 296* | L-18384 | 19 | 430 | L-3501 | 19 | 329 |
| AA2770 | 13 | 349 | ASD1887 | 22 | 282* | L-17672 | 20 | 430 | L-3344 | 54 | 328 |
| AA2895 | 58 | 316 | C0090 | 12 | 270 | L-17665 | 29 | 430 | D11 | 100 | 325 |
| AA0441 | 19 | 315 |  |  |  | L-20363 | 54 | 400 | L-3479 | 52 | 325 |
| AA2809 | 12 | 313 |  |  |  | L-15166 | 19 | 350 | L-3013 | 19 | 325 |
| AA2775 | 57 | 311 |  |  |  | L-15764 | 87 | 350 | A10 | 19 | 290 |
| AA2845 | 19 | 296 |  |  |  | L-14630 | 19 | 342 | C3 | 27 | 282 |
| AA2766 | 185 | 289 |  |  |  | L-10410 | 25 | 315 | A5 | 19 | 257 |
| AA2933 | 20 | 284 |  |  |  | L-15891 | 54 | 312 |  |  |  |
| AA2926 | 12 | 268 |  |  |  | L-15754 | 19 | 290 |  |  |  |
| AA2908 | 14 | 256 |  |  |  | L-15629 | 19 | 270 |  |  |  |
|  |  |  |  |  |  | L-15739 | 59 | 270 |  |  |  |
|  |  |  |  |  |  | L-15713 | 19 | 270 |  |  |  |

\*These samples have a (GAAGGA)<sub>n</sub> motif and are considered non-pathogenic.

**Table S3.**
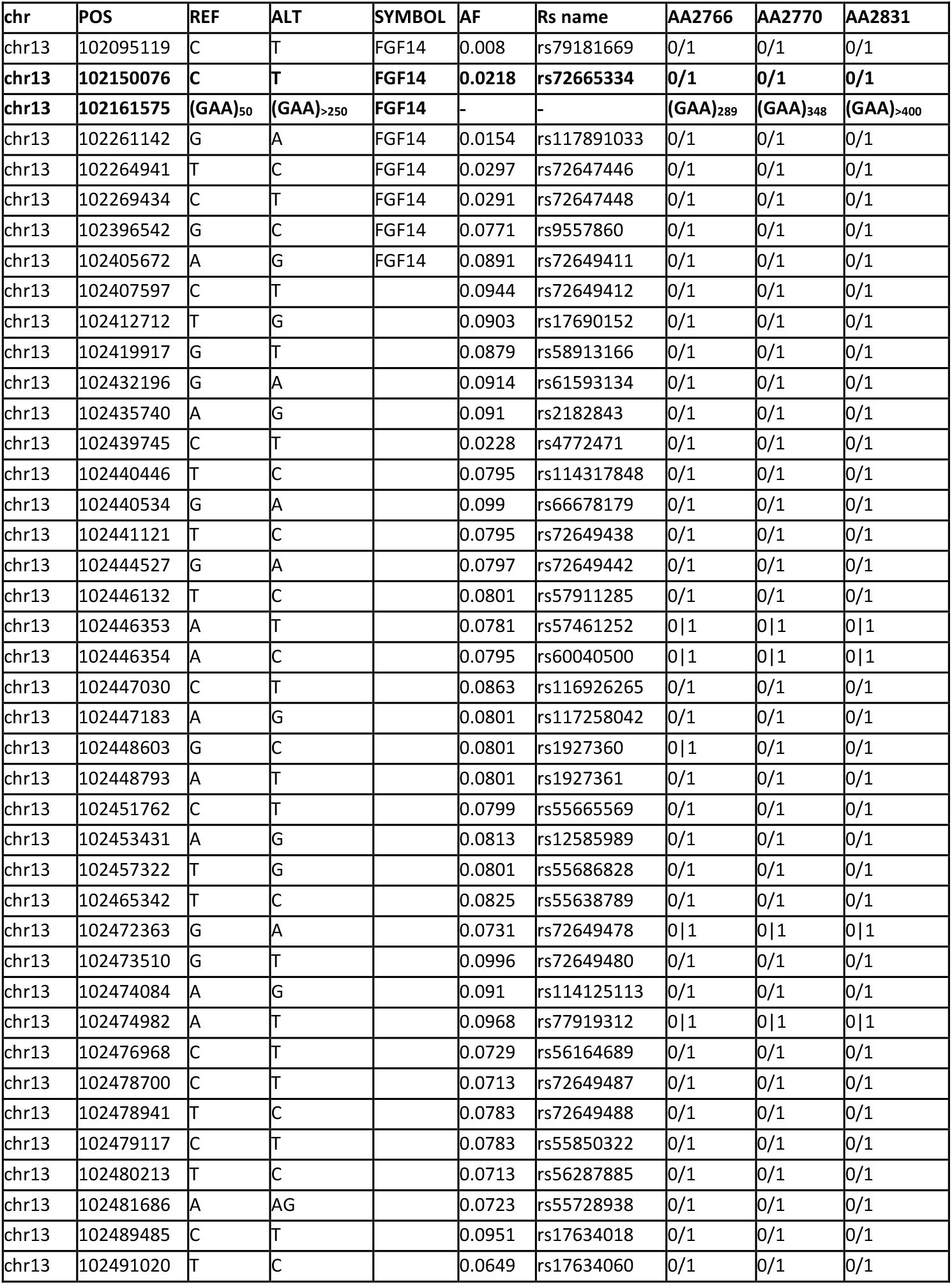
Summary of *FGF14* core haplotype shared by three Australian individuals with (GAA)_>250_.

| chr | POS | REF | ALT | SYMBOL | AF | Rs name | AA2766 | AA2770 | AA2831 |
| --- | --- | --- | --- | --- | --- | --- | --- | --- | --- |
| chr13 | 102095119 | C | T | FGF14 | 0.008 | rs79181669 | 0/1 | 0/1 | 0/1 |
| **chr13** | **102150076** | **C** | **T** | **FGF14** | **0.0218** | **rs72665334** | **0/1** | **0/1** | **0/1** |
| **chr13** | **102161575** | **(GAA)<sub>50</sub>** | **(GAA)<sub>>250</sub>** | **FGF14** | - | - | **(GAA)<sub>289</sub>** | **(GAA)<sub>348</sub>** | **(GAA)<sub>>400</sub>** |
| chr13 | 102261142 | G | A | FGF14 | 0.0154 | rs117891033 | 0/1 | 0/1 | 0/1 |
| chr13 | 102264941 | T | C | FGF14 | 0.0297 | rs72647446 | 0/1 | 0/1 | 0/1 |
| chr13 | 102269434 | C | T | FGF14 | 0.0291 | rs72647448 | 0/1 | 0/1 | 0/1 |
| chr13 | 102396542 | G | C | FGF14 | 0.0771 | rs9557860 | 0/1 | 0/1 | 0/1 |
| chr13 | 102405672 | A | G | FGF14 | 0.0891 | rs72649411 | 0/1 | 0/1 | 0/1 |
| chr13 | 102407597 | C | T |  | 0.0944 | rs72649412 | 0/1 | 0/1 | 0/1 |
| chr13 | 102412712 | T | G |  | 0.0903 | rs17690152 | 0/1 | 0/1 | 0/1 |
| chr13 | 102419917 | G | T |  | 0.0879 | rs58913166 | 0/1 | 0/1 | 0/1 |
| chr13 | 102432196 | G | A |  | 0.0914 | rs61593134 | 0/1 | 0/1 | 0/1 |
| chr13 | 102435740 | A | G |  | 0.091 | rs2182843 | 0/1 | 0/1 | 0/1 |
| chr13 | 102439745 | C | T |  | 0.0228 | rs4772471 | 0/1 | 0/1 | 0/1 |
| chr13 | 102440446 | T | C |  | 0.0795 | rs114317848 | 0/1 | 0/1 | 0/1 |
| chr13 | 102440534 | G | A |  | 0.099 | rs66678179 | 0/1 | 0/1 | 0/1 |
| chr13 | 102441121 | T | C |  | 0.0795 | rs72649438 | 0/1 | 0/1 | 0/1 |
| chr13 | 102444527 | G | A |  | 0.0797 | rs72649442 | 0/1 | 0/1 | 0/1 |
| chr13 | 102446132 | T | C |  | 0.0801 | rs57911285 | 0/1 | 0/1 | 0/1 |
| chr13 | 102446353 | A | T |  | 0.0781 | rs57461252 | 0\|1 | 0\|1 | 0\|1 |
| chr13 | 102446354 | A | C |  | 0.0795 | rs60040500 | 0\|1 | 0\|1 | 0\|1 |
| chr13 | 102447030 | C | T |  | 0.0863 | rs116926265 | 0/1 | 0/1 | 0/1 |
| chr13 | 102447183 | A | G |  | 0.0801 | rs117258042 | 0/1 | 0/1 | 0/1 |
| chr13 | 102448603 | G | C |  | 0.0801 | rs1927360 | 0\|1 | 0/1 | 0/1 |
| chr13 | 102448793 | A | T |  | 0.0801 | rs1927361 | 0/1 | 0/1 | 0/1 |
| chr13 | 102451762 | C | T |  | 0.0799 | rs55665569 | 0/1 | 0/1 | 0/1 |
| chr13 | 102453431 | A | G |  | 0.0813 | rs12585989 | 0/1 | 0/1 | 0/1 |
| chr13 | 102457322 | T | G |  | 0.0801 | rs55686828 | 0/1 | 0/1 | 0/1 |
| chr13 | 102465342 | T | C |  | 0.0825 | rs55638789 | 0/1 | 0/1 | 0/1 |
| chr13 | 102472363 | G | A |  | 0.0731 | rs72649478 | 0\|1 | 0\|1 | 0\|1 |
| chr13 | 102473510 | G | T |  | 0.0996 | rs72649480 | 0/1 | 0/1 | 0/1 |
| chr13 | 102474084 | A | G |  | 0.091 | rs114125113 | 0/1 | 0/1 | 0/1 |
| chr13 | 102474982 | A | T |  | 0.0968 | rs77919312 | 0\|1 | 0\|1 | 0\|1 |
| chr13 | 102476968 | C | T |  | 0.0729 | rs56164689 | 0/1 | 0/1 | 0/1 |
| chr13 | 102478700 | C | T |  | 0.0713 | rs72649487 | 0/1 | 0/1 | 0/1 |
| chr13 | 102478941 | T | C |  | 0.0783 | rs72649488 | 0/1 | 0/1 | 0/1 |
| chr13 | 102479117 | C | T |  | 0.0783 | rs55850322 | 0/1 | 0/1 | 0/1 |
| chr13 | 102480213 | T | C |  | 0.0713 | rs56287885 | 0/1 | 0/1 | 0/1 |
| chr13 | 102481686 | A | AG |  | 0.0723 | rs55728938 | 0/1 | 0/1 | 0/1 |
| chr13 | 102489485 | C | T |  | 0.0951 | rs17634018 | 0/1 | 0/1 | 0/1 |
| chr13 | 102491020 | T | C |  | 0.0649 | rs17634060 | 0/1 | 0/1 | 0/1 |

## Notes

### Competing Interest Statement

The authors have declared no competing interest.

### Author Declarations

The Royal Childrens Hospital Human Research Ethics Committee approved this work (HREC 28097). The Walter and Eliza Hall Institute of Medical Research Ethics Committee approved this work (HREC 18/06). The Ethics committee of the University of Lubeck approved this work (AZ16-039).

## REFERENCES

1. Ruano, L., Melo, C., Silva, M.C., and Coutinho, P. (2014). The global epidemiology of hereditary ataxia and spastic paraplegia: a systematic review of prevalence studies. Neuroepidemiology 42, 174-183. 10.1159/000358801.

2. Benson, G. (1999). Tandem repeats finder: a program to analyze DNA sequences. Nucleic Acids Res 27, 573–580.

3. Mousavi, N., Shleizer-Burko, S., Yanicky, R., and Gymrek, M. (2019). Profiling the genome-wide landscape of tandem repeat expansions. Nucleic Acids Res 47, e90. 10.1093/nar/gkz501.

4. Depienne, C., and Mandel, J.L. (2021). 30 years of repeat expansion disorders: What have we learned and what are the remaining challenges? American journal of human genetics 108, 764-785. 10.1016/j.ajhg.2021.03.011.

5. Harding, A.E. (1981). “Idiopathic” late onset cerebellar ataxia. A clinical and genetic study of 36 cases. J Neurol Sci 51, 259–271.

6. Klockgether, T., Kramer, B., Ludtke, R., Schols, L., and Laccone, F. (1996). Repeat length and disease progression in spinocerebellar ataxia type 3. Lancet 348, 830. 10.1016/S0140-6736(05)65255-5.

7. Rexach, J., Lee, H., Martinez-Agosto, J.A., Nemeth, A.H., and Fogel, B.L. (2019). Clinical application of next-generation sequencing to the practice of neurology. Lancet Neurol 18, 492–503. 10.1016/S1474-4422(19)30033-X.

8. Bahlo, M., Bennett, M.F., Degorski, P., Tankard, R.M., Delatycki, M.B., and Lockhart, P.J. (2018). Recent advances in the detection of repeat expansions with short-read next-generation sequencing. F1000Res 7. 10.12688/f1000research.13980.1.

9. Dolzhenko, E., Bennett, M.F., Richmond, P.A., Trost, B., Chen, S., van Vugt, J., Nguyen, C., Narzisi, G., Gainullin, V.G., Gross, A.M., et al. (2020). ExpansionHunter Denovo: a computational method for locating known and novel repeat expansions in short-read sequencing data. Genome biology 21, 102. 10.1186/s13059-020-02017-z.

10. Rafehi, H., Szmulewicz, D.J., Bennett, M.F., Sobreira, N.L.M., Pope, K., Smith, K.R., Gillies, G., Diakumis, P., Dolzhenko, E., Eberle, M.A., et al. (2019). Bioinformatics-Based Identification of Expanded Repeats: A Non-reference Intronic Pentamer Expansion in RFC1 Causes CANVAS. American journal of human genetics 105, 151–165. 10.1016/j.ajhg.2019.05.016.

11. Payne, A., Holmes, N., Clarke, T., Munro, R., Debebe, B.J., and Loose, M. (2021). Readfish enables targeted nanopore sequencing of gigabase-sized genomes. Nature biotechnology 39, 442–450. 10.1038/s41587-020-00746-x.

12. DePristo, M.A., Banks, E., Poplin, R., Garimella, K.V., Maguire, J.R., Hartl, C., Philippakis, A.A., del Angel, G., Rivas, M.A., Hanna, M., et al. (2011). A framework for variation discovery and genotyping using next-generation DNA sequencing data. Nature genetics 43, 491–498. 10.1038/ng.806.

13. Van der Auwera, G.A., Carneiro, M.O., Hartl, C., Poplin, R., Del Angel, G., Levy-Moonshine, A., Jordan, T., Shakir, K., Roazen, D., Thibault, J., et al. (2013). From FastQ data to high confidence variant calls: the Genome Analysis Toolkit best practices pipeline. Curr Protoc Bioinformatics 43, 11 10 11–11 10 33. 10.1002/0471250953.bi1110s43.

14. Tankard, R.M., Bennett, M.F., Degorski, P., Delatycki, M.B., Lockhart, P.J., and Bahlo, M. (2018). Detecting Expansions of Tandem Repeats in Cohorts Sequenced with Short-Read Sequencing Data. American journal of human genetics 103, 858–873. 10.1016/j.ajhg.2018.10.015.

15. Dolzhenko, E., Deshpande, V., Schlesinger, F., Krusche, P., Petrovski, R., Chen, S., Emig-Agius, D., Gross, A., Narzisi, G., Bowman, B., et al. (2019). ExpansionHunter: a sequence-graph-based tool to analyze variation in short tandem repeat regions. Bioinformatics 35, 4754–4756. 10.1093/bioinformatics/btz431.

16. Genomes Project, C., Auton, A., Brooks, L.D., Durbin, R.M., Garrison, E.P., Kang, H.M., Korbel, J.O., Marchini, J.L., McCarthy, S., McVean, G.A., and Abecasis, G.R. (2015). A global reference for human genetic variation. Nature 526, 68–74. 10.1038/nature15393.

17. Byrska-Bishop, M., Evani, U.S., Zhao, X., Basile, A.O., Abel, H.J., Regier, A.A., Corvelo, A., Clarke, W.E., Musunuri, R., Nagulapalli, K., et al. (2022). High-coverage whole-genome sequencing of the expanded 1000 Genomes Project cohort including 602 trios. Cell 185, 3426–3440 e3419. 10.1016/j.cell.2022.08.004.

18. Di Tommaso, P., Chatzou, M., Floden, E.W., Barja, P.P., Palumbo, E., and Notredame, C. (2017). Nextflow enables reproducible computational workflows. Nature biotechnology 35, 316–319. 10.1038/nbt.3820.

19. McKenna, A., Hanna, M., Banks, E., Sivachenko, A., Cibulskis, K., Kernytsky, A., Garimella, K., Altshuler, D., Gabriel, S., Daly, M., and DePristo, M.A. (2010). The Genome Analysis Toolkit: a MapReduce framework for analyzing next-generation DNA sequencing data. Genome research 20, 1297–1303. 10.1101/gr.107524.110.

20. Li, H. (2011). A statistical framework for SNP calling, mutation discovery, association mapping and population genetical parameter estimation from sequencing data. Bioinformatics 27, 2987–2993. 10.1093/bioinformatics/btr509.

21. Robinson, J.T., Thorvaldsdottir, H., Winckler, W., Guttman, M., Lander, E.S., Getz, G., and Mesirov, J.P. (2011). Integrative genomics viewer. Nature biotechnology 29, 24–26. 10.1038/nbt.1754.

22. McLaren, W., Gil, L., Hunt, S.E., Riat, H.S., Ritchie, G.R., Thormann, A., Flicek, P., and Cunningham, F. (2016). The Ensembl Variant Effect Predictor. Genome biology 17, 122. 10.1186/s13059-016-0974-4.

23. Mitsuhashi, S., Frith, M.C., Mizuguchi, T., Miyatake, S., Toyota, T., Adachi, H., Oma, Y., Kino, Y., Mitsuhashi, H., and Matsumoto, N. (2019). Tandem-genotypes: robust detection of tandem repeat expansions from long DNA reads. Genome biology 20, 58. 10.1186/s13059-019-1667-6.

24. Wilson, G.R., Sunley, J., Smith, K.R., Pope, K., Bromhead, C.J., Fitzpatrick, E., Di Rocco, M., van Steensel, M., Coman, D.J., Leventer, R.J., et al. (2013). Mutations in SH3PXD2B cause Borrone dermato-cardio-skeletal syndrome. European journal of human genetics : EJHG. 10.1038/ejhg.2013.229.

25. Barbier, M., Bahlo, M., Pennisi, A., Jacoupy, M., Tankard, R.M., Ewenczyk, C., Davies, K.C., Lino-Coulon, P., Colace, C., Rafehi, H., et al. (2022). Heterozygous PNPT1 Variants Cause Spinocerebellar Ataxia Type 25. Annals of neurology 92, 122–137. 10.1002/ana.26366.

26. van Swieten, J.C., Brusse, E., de Graaf, B.M., Krieger, E., van de Graaf, R., de Koning, I., Maat-Kievit, A., Leegwater, P., Dooijes, D., Oostra, B.A., and Heutink, P. (2003). A mutation in the fibroblast growth factor 14 gene is associated with autosomal dominant cerebellar ataxia [corrected]. American journal of human genetics 72, 191–199. 10.1086/345488.

27. Zuhlke, C., Dalski, A., Hellenbroich, Y., Bubel, S., Schwinger, E., and Burk, K. (2002). Spinocerebellar ataxia type 1 (SCA1): phenotype-genotype correlation studies in intermediate alleles. European journal of human genetics : EJHG 10, 204–209. 10.1038/sj.ejhg.5200788.

28. Menon, R.P., Nethisinghe, S., Faggiano, S., Vannocci, T., Rezaei, H., Pemble, S., Sweeney, M.G., Wood, N.W., Davis, M.B., Pastore, A., and Giunti, P. (2013). The role of interruptions in polyQ in the pathology of SCA1. PLoS Genet 9, e1003648. 10.1371/journal.pgen.1003648.

29. Pearson, C.E., Eichler, E.E., Lorenzetti, D., Kramer, S.F., Zoghbi, H.Y., Nelson, D.L., and Sinden, R.R. (1998). Interruptions in the triplet repeats of SCA1 and FRAXA reduce the propensity and complexity of slipped strand DNA (S-DNA) formation. Biochemistry 37, 2701–2708. 10.1021/bi972546c.

30. Wang, Q., McEwen, D.G., and Ornitz, D.M. (2000). Subcellular and developmental expression of alternatively spliced forms of fibroblast growth factor 14. Mech Dev 90, 283–287. 10.1016/s0925-4773(99)00241-5.

31. Bidichandani, S.I., Ashizawa, T., and Patel, P.I. (1998). The GAA triplet-repeat expansion in Friedreich ataxia interferes with transcription and may be associated with an unusual DNA structure. American journal of human genetics 62, 111–121. 10.1086/301680.

32. Kuhlmann, K., Cieselski, M., and Schumann, J. (2021). Relative versus absolute RNA quantification: a comparative analysis based on the example of endothelial expression of vasoactive receptors. Biol Proced Online 23, 6. 10.1186/s12575-021-00144-w.

33. MacDougall, H.G., Weber, K.P., McGarvie, L.A., Halmagyi, G.M., and Curthoys, I.S. (2009). The video head impulse test: diagnostic accuracy in peripheral vestibulopathy. Neurology 73, 1134–1141. 10.1212/WNL.0b013e3181bacf85.

34. Szmulewicz, D., MacDougall, H., Storey, E., Curthoys, I., and Halmagyi, M. (2014). A Novel Quantitative Bedside Test of Balance Function: The Video Visually Enhanced Vestibulo-ocular Reflex (VVOR) Neurology 82, S19.002.

35. Paulson, H. (2018). Repeat expansion diseases. Handbook of clinical neurology 147, 105–123. 10.1016/B978-0-444-63233-3.00009-9.

36. Ibanez, K., Polke, J., Hagelstrom, R.T., Dolzhenko, E., Pasko, D., Thomas, E.R.A., Daugherty, L.C., Kasperaviciute, D., Smith, K.R., Group, W.G.S.f.N.D., et al. (2022). Whole genome sequencing for the diagnosis of neurological repeat expansion disorders in the UK: a retrospective diagnostic accuracy and prospective clinical validation study. Lancet Neurol 21, 234–245. 10.1016/S1474-4422(21)00462-2.

37. Strupp, M., Maul, S., Konte, B., Hartmann, A.M., Giegling, I., Wollenteit, S., Feil, K., and Rujescu, D. (2020). A Variation in FGF14 Is Associated with Downbeat Nystagmus in a Genome-Wide Association Study. Cerebellum 19, 348–357. 10.1007/s12311-020-01113-x.

38. Wagner, J.N., Glaser, M., Brandt, T., and Strupp, M. (2008). Downbeat nystagmus: aetiology and comorbidity in 117 patients. J Neurol Neurosurg Psychiatry 79, 672–677. 10.1136/jnnp.2007.126284.

39. Lou, J.Y., Laezza, F., Gerber, B.R., Xiao, M., Yamada, K.A., Hartmann, H., Craig, A.M., Nerbonne, J.M., and Ornitz, D.M. (2005). Fibroblast growth factor 14 is an intracellular modulator of voltage-gated sodium channels. J Physiol 569, 179–193. 10.1113/jphysiol.2005.097220.

40. Di Re, J., Wadsworth, P.A., and Laezza, F. (2017). Intracellular Fibroblast Growth Factor 14: Emerging Risk Factor for Brain Disorders. Front Cell Neurosci 11, 103. 10.3389/fncel.2017.00103.

41. Wang, Q., Bardgett, M.E., Wong, M., Wozniak, D.F., Lou, J., McNeil, B.D., Chen, C., Nardi, A., Reid, D.C., Yamada, K., and Ornitz, D.M. (2002). Ataxia and paroxysmal dyskinesia in mice lacking axonally transported FGF14. Neuron 35, 25–38. 10.1016/s0896-6273(02)00744-4.

42. Bosch, M.K., Carrasquillo, Y., Ransdell, J.L., Kanakamedala, A., Ornitz, D.M., and Nerbonne, J.M. (2015). Intracellular FGF14 (iFGF14) Is Required for Spontaneous and Evoked Firing in Cerebellar Purkinje Neurons and for Motor Coordination and Balance. J Neurosci 35, 6752–6769. 10.1523/JNEUROSCI.2663-14.2015.

43. Yan, H., Pablo, J.L., Wang, C., and Pitt, G.S. (2014). FGF14 modulates resurgent sodium current in mouse cerebellar Purkinje neurons. Elife 3, e04193. 10.7554/eLife.04193.

44. Sakamoto, N., Larson, J.E., Iyer, R.R., Montermini, L., Pandolfo, M., and Wells, R.D. (2001). GGA*TCC-interrupted triplets in long GAA*TTC repeats inhibit the formation of triplex and sticky DNA structures, alleviate transcription inhibition, and reduce genetic instabilities. The Journal of biological chemistry 276, 27178–27187. 10.1074/jbc.M101852200.

45. Misceo, D., Fannemel, M., Baroy, T., Roberto, R., Tvedt, B., Jaeger, T., Bryn, V., Stromme, P., and Frengen, E. (2009). SCA27 caused by a chromosome translocation: further delineation of the phenotype. Neurogenetics 10, 371–374. 10.1007/s10048-009-0197-x.

46. Groth, C.L., and Berman, B.D. (2018). Spinocerebellar Ataxia 27: A Review and Characterization of an Evolving Phenotype. Tremor Other Hyperkinet Mov (N Y) 8, 534. 10.7916/D80S0ZJQ.

47. Dalski, A., Atici, J., Kreuz, F.R., Hellenbroich, Y., Schwinger, E., and Zuhlke, C. (2005). Mutation analysis in the fibroblast growth factor 14 gene: frameshift mutation and polymorphisms in patients with inherited ataxias. European journal of human genetics : EJHG 13, 118–120. 10.1038/sj.ejhg.5201286.

48. Miura, S., Kosaka, K., Fujioka, R., Uchiyama, Y., Shimojo, T., Morikawa, T., Irie, A., Taniwaki, T., and Shibata, H. (2019). Spinocerebellar ataxia 27 with a novel nonsense variant (Lys177X) in FGF14. Eur J Med Genet 62, 172–176. 10.1016/j.ejmg.2018.07.005.

49. Paucar, M., Lundin, J., Alshammari, T., Bergendal, A., Lindefeldt, M., Alshammari, M., Solders, G., Di Re, J., Savitcheva, I., Granberg, T., et al. (2020). Broader phenotypic traits and widespread brain hypometabolism in spinocerebellar ataxia 27. J Intern Med 288, 103–115. 10.1111/joim.13052.

50. Casey, H.L., and Gomez, C.M. (1993). Spinocerebellar Ataxia Type 6. In GeneReviews((R)), M.P. Adam, D.B. Everman, G.M. Mirzaa, R.A. Pagon, S.E. Wallace, L.J.H. Bean, K.W. Gripp, and A. Amemiya, eds.

51. Gisatulin, M., Dobricic, V., Zühlke, C., Hellenbroich, Y., Tadic, V., Münchau, A., Isenhardt, K., Bürk, K., Bahlo, M., Lockhart, P.J., et al. (2020). Clinical spectrum of the pentanucleotide repeat expansion in the RFC1 gene in ataxia syndromes. Neurology Accepted 30th May.

52. Montaut, S., Diedhiou, N., Fahrer, P., Marelli, C., Lhermitte, B., Robelin, L., Vincent, M.C., Corti, L., Taieb, G., Gebus, O., et al. (2021). Biallelic RFC1-expansion in a French multicentric sporadic ataxia cohort. J Neurol 268, 3337–3343. 10.1007/s00415-021-10499-5.

53. Davies, K., Szmulewicz, D.J., Corben, L.A., Delatycki, M., and Lockhart, P.J. (2022). RFC1-Related Disease: Molecular and Clinical Insights. Neurol Genet 8, e200016. 10.1212/NXG.0000000000200016.

54. Tezenas du Montcel, S., Durr, A., Bauer, P., Figueroa, K.P., Ichikawa, Y., Brussino, A., Forlani, S., Rakowicz, M., Schols, L., Mariotti, C., et al. (2014). Modulation of the age at onset in spinocerebellar ataxia by CAG tracts in various genes. Brain 137, 2444–2455. 10.1093/brain/awu174.

55. Delatycki, M.B., Williamson, R., and Forrest, S.M. (2000). Friedreich ataxia: an overview. Journal of medical genetics 37, 1–8. 10.1136/jmg.37.1.1.

56. Lange, L.M., Gonzalez-Latapi, P., Rajalingam, R., Tijssen, M.A.J., Ebrahimi-Fakhari, D., Gabbert, C., Ganos, C., Ghosh, R., Kumar, K.R., Lang, A.E., et al. (2022). Nomenclature of Genetic Movement Disorders: Recommendations of the International Parkinson and Movement Disorder Society Task Force - An Update. Mov Disord 37, 905–935. 10.1002/mds.28982.

57. Marras, C., Lang, A., van de Warrenburg, B.P., Sue, C.M., Tabrizi, S.J., Bertram, L., Mercimek-Mahmutoglu, S., Ebrahimi-Fakhari, D., Warner, T.T., Durr, A., et al. (2016). Nomenclature of genetic movement disorders: Recommendations of the international Parkinson and movement disorder society task force. Mov Disord 31, 436–457. 10.1002/mds.26527.

58. Lockhart, P.J. (2022). Advancing the diagnosis of repeat expansion disorders. Lancet Neurol 21, 205–207. 10.1016/S1474-4422(22)00033-3.

